# Estimating the burden of RSV- and influenza-associated hospitalizations, ICU admissions, and deaths across age and socioeconomic groups in New York State, 2005-2019

**DOI:** 10.1101/2025.01.10.24319265

**Authors:** Hanmeng Xu, Virginia E. Pitzer, Joshua L. Warren, Eugene D. Shapiro, Daniel M. Weinberger

## Abstract

**Background:** Multiple prophylactic products are now available to protect against respiratory syncytial virus (RSV) in different age groups. Assessing the pre-intervention burden of RSV infections across various severity levels and risk groups is crucial, as it provides a baseline for evaluating the impact of these products.

**Methods:** We obtained monthly time series data on hospitalizations, intensive care unit (ICU) admissions, and deaths by age group, ZIP code, and cause for New York state from 2005 to 2019. Socioeconomic status (SES) of the ZIP codes was classified using supervised principal component analysis (PCA). We estimated the incidence of hospitalizations, ICU admissions, and deaths attributable to RSV and to influenza using hierarchical Bayesian regression models. Additionally, we assessed severity, defined by ICU admission and mortality risks, as well as recording fraction (i.e., percent of estimated virus-associated hospitalizations recorded as being due to the specific virus), stratified by age, SES, and over time.

**Results:** The estimated annual incidence of RSV-associated hospitalizations and ICU admissions were highest in infants under 1 in the low SES group (2,240 [95% credible interval (CrI): 2,200-2,290] hospitalizations and 330 [95% CrI: 320-350] ICU admissions per 100,000 person-years). The incidence of RSV-associated deaths was highest among adults ≥85 years old (61 [95% CrI: 49-74] per 100,000 person-years). In contrast to RSV, the burden of influenza was greatest in age groups ≥65 years. The risk of ICU admission varied by patients’ age and SES, and the mortality risk increased dramatically with age for both pathogens (RSV: 11.9% [95% CrI: 9.6-14.3%], influenza: 14.4% [95% CrI: 13.1-15.6%] among ≥85 year age group). Incidence varied by epidemic year and season, and we observed an increasing recording fraction of RSV among all age groups over the study period.

**Conclusions:** RSV and influenza contribute significantly to the burden of hospitalizations, ICU admissions, and deaths, particularly among infants and older adults. Although the recording fraction of RSV increased over the study period, it remains lower, particularly for adults. Our findings reveal a disparity in hospitalization burden by SES, particularly among younger age groups.

## Introduction

Respiratory illnesses caused by viral infections impose a substantial burden of hospitalizations, ICU admissions, and mortality [1]. Respiratory syncytial virus (RSV) and influenza are among the most common pathogens causing lower respiratory tract infections (LRTI), with a particularly high burden in infants, young children, and the elderly [1–3]. Globally, RSV is estimated to cause approximately 3.6 million acute LRTI hospitalizations and 26,300 in-hospital deaths among children aged 0-60 months annually [2]. Comparably, the World Health Organization estimated that globally 3-5 million cases of severe illness and 290,000-650,000 deaths are caused by seasonal influenza per year [4]. Underlying health conditions (e.g., prematurity, chronic lung diseases, congenital heart diseases, compromised immune system) and socioeconomic conditions could increase susceptibility to RSV and influenza infection and severe disease [4–7].

Several prophylactic products protecting against RSV have been approved in the US since 2023, including three vaccines for adults aged 60 years and older (one of which was also approved for pregnant women to protect their newborns) [8,9] and the first long-acting monoclonal antibody (nirsevimab) for infants [10][9]. Evaluating the potential impact of these novel preventive interventions requires understanding the baseline burden of disease in different risk groups and for different outcomes.

Estimation of the burden of RSV and influenza is complicated by patterns of healthcare seeking, diagnostic practices, and reporting standards of different healthcare systems. Previous work estimated that the incidence of RSV-associated hospitalizations was highest among infants under 1, followed by adults 85 years of age and older, and was higher among individuals from low-income ZIP codes [11], while the burden of Influenza-associated hospitalizations was estimated to be highest among older adults [12]. In terms of more severe outcomes, i.e., intensive care unit (ICU) admission and death, higher incidence was found among older adults for both pathogens [3,13–15]. Given the variation in RSV and influenza burden across different clinical outcomes, age groups, and SES groups, a more comprehensive assessment is needed.

This study aimed to quantify the annual incidence of hospitalizations, ICU admissions, and deaths due to RSV infection across different age and SES groups, and to contextualize the findings by comparing them to the estimated disease burden attributable to influenza. Our estimates of RSV burden for different subgroups can also serve as a pre-intervention baseline to assess the impact of recently approved prophylactic interventions.

## Methods

### Data sources

Individual-level hospital discharge data from New York state prior to the COVID-19 pandemic from July 2005 to June 2019 were obtained from the State Inpatient Databases of the Healthcare Cost and Utilization Project (HCUP) [16]. As the coding system of the International Classification of Diseases (ICD) shifted from ICD-9-CM to ICD-10-CM in 2015, data from 2015 were not included in the analysis. Variables extracted from the database included age at admission, ZIP code of residence, ICD-9-CM (2005-2014) or ICD-10-CM diagnostic codes (2016-2019), and a binary indicator for death during hospitalization. We identified ICU admissions by reviewing individual charge data and marking those with ICU-related charges as individuals admitted to ICU, specifically those with UB-92 revenue center codes 200-219 [17]. We used the ICD-9-CM codes 079.6, 466.11, 480.1 and ICD-10-CM codes B97.4, J21.0, J12.1, J20.5 to identify RSV-coded hospitalizations, and ICD-9-CM codes 487 and ICD-10-CM codes J09, J10, J11 to identify influenza-coded hospitalizations and ICD-9-CM codes 460-519 and ICD-10-CM codes J00-J99 to identify all-cause respiratory hospitalizations.

The population was stratified into nine age groups: <1, 1, 2-4, 5-9, 10-19, 20-44, 45-64, 65-84, and ≥85 years old. Population size estimates for New York state by age and ZIP code were obtained from the CDC WONDER online database [18]. We classified ZIP codes into three SES groups (low, medium, and high) using supervised principal component analysis (PCA) in which the incidence of RSV-coded hospitalization at the ZIP code level was the target variable (see supplementary methods, Figure S1-2)[19]. The first principal component was derived from seven economic indicators at the ZIP code level, including household income, employment rate, household size, population density, and racial component. The resulting factor scores were used to categorize all the ZIP codes into three equal-sized SES tertiles. Patients in our dataset were assigned to one of the three SES groups based on their recorded residential ZIP code, as individual-level indicators of SES are not available.

The individual-level data were aggregated into monthly time series by age and SES group, providing for each month the count of all-cause respiratory hospitalizations, ICU admissions, and deaths. We also created time series for RSV-coded hospitalizations under 2 years old by SES and influenza-coded hospitalizations among all age groups by SES, which we used as indicators of RSV and influenza infections over time in the model.

### Statistical model

We used hierarchical Bayesian Poisson regression models to estimate the incidence of all-cause respiratory hospitalizations, ICU admissions, and deaths that could be attributed to RSV and influenza. The analyses were stratified by age and SES groups. The expected number of all-cause respiratory hospitalizations in age group *j* and SES group *k* in month *i, λ*_*ijk*_, was modeled as a function of RSV-coded hospitalizations (under 2 years old), influenza-coded hospitalizations (among all ages), baseline seasonal variation, and temporal trends. The effects of each predictor were assumed to be additive. All-cause ICU admissions and deaths were modeled similarly.

The three clinical outcomes were linked in the model through disease progression risks, with the rationale that in-hospital ICU admissions and deaths are subsets of hospitalizations. We used weakly informative priors for the parameters in our model. A full description of the model structure and methods to summarize model outputs can be found in the supplementary methods. Data cleaning of the HCUP data was performed with SAS software, version 9.2 (SAS Institute, Cary, North Carolina), and R version 4.3.1 [20,21]. Statistical analyses were performed in R version 4.3.1 [21]. The model code is available on GitHub (https://github.com/DanWeinberger/rsv_burden_deaths_us).

## Results

### Recorded hospitalizations, ICU admissions, and deaths

From 2005/06 to 2018/19 (total 12 seasons due to the ICD transition in 2015), an annual average of 6200 hospitalizations, 1100 ICU admissions, and 63 deaths were recorded as being due to RSV. Among these events attributed to RSV, 55.4% of hospitalizations, 56.1% of ICU admissions, and 5.4% of deaths occurred in children <1 year old and 10.9% of hospitalizations, 9.8% of ICU admissions and 63.0% of deaths occurred in people 65 years old and above. In contrast, an average of 7900 hospitalizations, 1200 ICU admissions, and 250 deaths were recorded as due to influenza each season. Of these influenza-attributed events, 5.1% of hospitalizations, 4.4% of ICU admissions, and 0.4% of deaths occurred in children <1, and 44.5% of hospitalizations, 42.3% of ICU admissions, and 69.1% of deaths occurred in people 65 years and above. The annual incidence of RSV-coded hospitalizations and ICU admissions was substantially higher among children under 5 (Figure S4). Older adults had a higher incidence of both RSV- and influenza-coded deaths.

### Estimated incidence of RSV-associated disease outcomes

Based on our statistical model, the estimated incidence of RSV-associated hospitalizations and ICU admissions followed a U-shaped pattern, with the highest rates in the youngest and oldest age groups (Figure 1). The estimated incidence rate was highest in infants <1 year, at 1920 (95% credible interval (CrI): 1900-1950) per 100,000 person-years, followed by children aged 1-<2 years (600, 95% CrI: 580-610) and adults ≥85 years (510, 95% CrI: 480-540).

**Figure 1.**
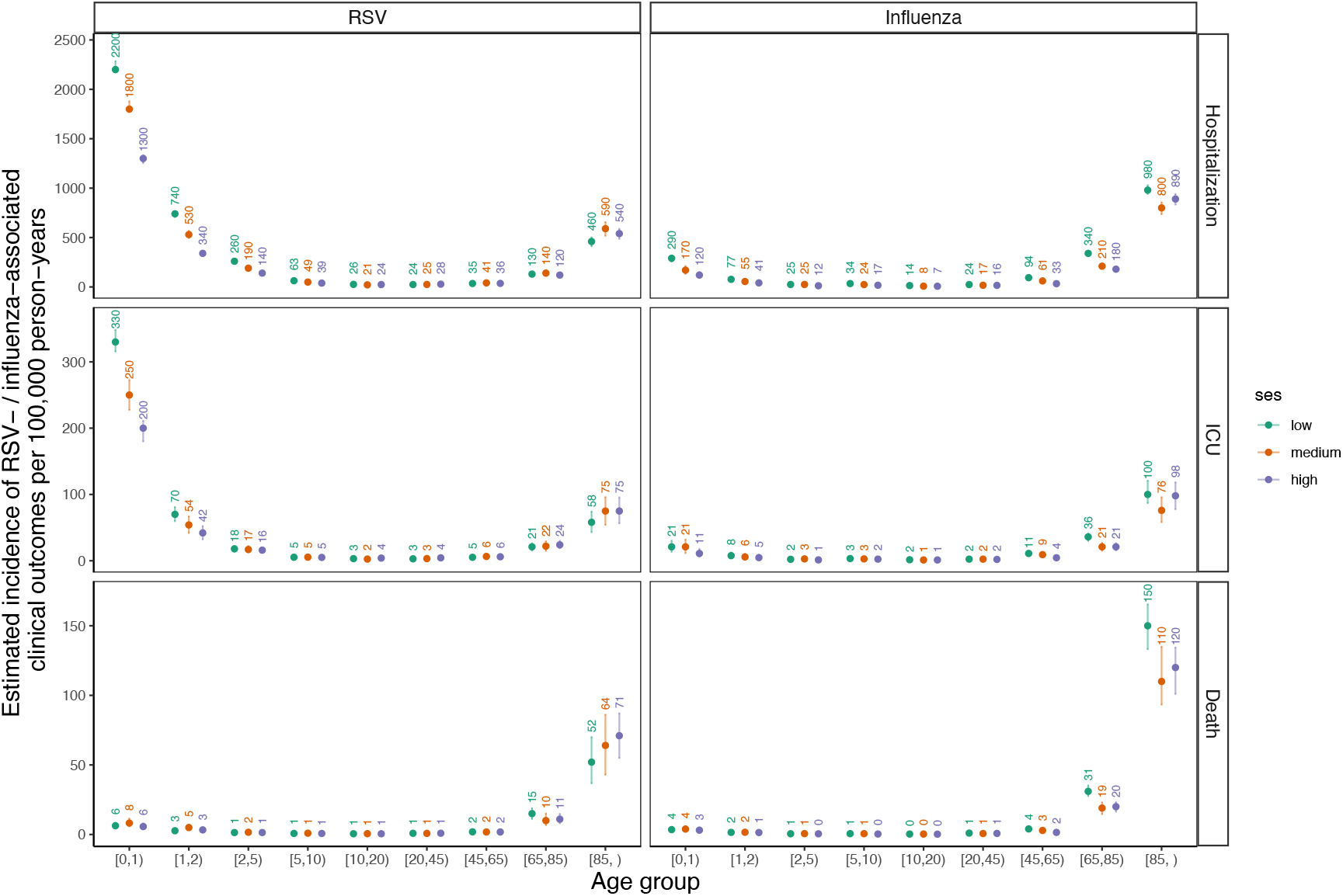
Estimated incidence of RSV- and influenza-associated hospitalizations (A), ICU admissions (B), and deaths (C), 2005-2019. The dots indicate the posterior median estimates of the incidence of hospitalizations, ICU admissions, and deaths that are attributable to RSV or influenza per 100,000 person-years, with the labels indicating the estimated values. The error bars indicate the 95% credible intervals of the estimated incidence.

The incidence of RSV-associated ICU admissions showed a similar U-shape as hospitalizations, but the incidence of RSV-associated deaths displayed a J-shaped distribution by age (Figure 1), with a much higher incidence in the age groups 65 years and older. Overall, around 55% of RSV-associated hospitalizations and 61% of RSV-associated ICU admissions occurred in children under 1, while 83% of RSV-associated deaths occurred in adults aged 65 and above.

Higher incidence of RSV hospitalizations and ICU admissions were observed in the low SES group among children under 10 but not other ages (Figure 1). In infants <1 year, the incidence rate of RSV hospitalizations was nearly twice as high in the low SES group (2200 per 100,000 person-years) compared to the high SES group (1300 per 100,000 person-years). However, there were no clear differences in the rate of death from RSV by SES group.

In contrast to RSV, incidence rate estimates for all three influenza-associated outcomes followed a J-shape distribution by age, with the highest burden in the oldest age groups 65 years and older (Figure 1). The incidence of influenza-associated hospitalization and ICU admission was markedly lower for children compared with RSV (e.g., 220 [95%CrI: 200-240] vs 1920 [95%CrI: 1900-1950] hospitalizations per 100,000 person-years for infants <1 year of age, unstratified by SES group), while the incidence rate for influenza-associated outcomes was higher than for RSV among older adults 65 years and older, especially for deaths. Overall, about 72% of influenza-associated hospitalizations, 74% of influenza-associated ICU admissions, and 93% of influenza-associated deaths occurred in adults aged 65 and above.

### Disease burden over time

The burden of disease estimated to be caused by RSV and influenza varied over time (Figure 2). We estimated a relatively stable trend with a slight decrease in RSV-associated hospitalizations among infants <1 year and a decreasing trend in all three RSV-associated outcomes among adults 65 years and older, in contrast to the increase in RSV-coded outcomes (Figure 2-4). However, ICU admissions almost doubled in infants <1 year between 2013/14 and 2018/19. This rise in ICU admissions for infants coincided with shorter hospital stays, increased use of non-invasive mechanical ventilation (NIMV), and decreased use of invasive mechanical ventilation (IMV) (Figures S7, S8). A larger decrease in the length of stay in the hospital also coincided with a larger increase in ICU admissions in the low SES group (Figure S7, S9). In contrast with RSV-associated burden over time, influenza-associated outcomes remained relatively stable over the study period, albeit with more season-to-season variability for older age groups (Figure 2, S5, S6).

**Figure 2.**
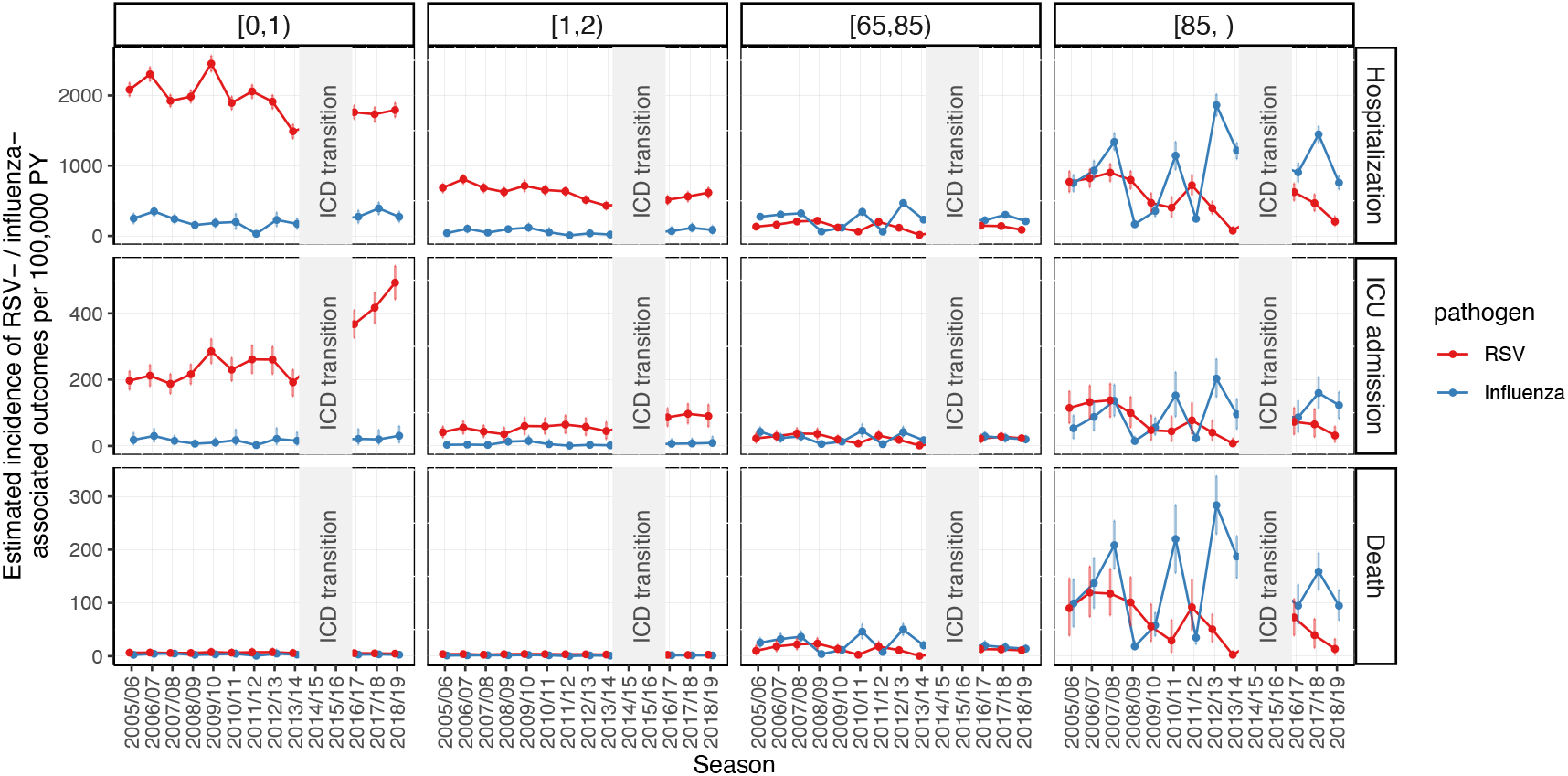
Estimated incidence of RSV- and influenza-associated hospitalizations, ICU admissions, and deaths over time by age group, 2005-2019. The pale grey blocks represent the time period of transition of the ICD coding system from ICD-9-CM to ICD-10-CM in 2015. The dots represent the posterior median estimates of the incidence rate of hospitalizations, ICU admissions, and deaths that are attributable to RSV or influenza per 100,000 person-years for each season. The error bars indicate the 95% credible intervals of the estimated incidence. The four vertical panels indicate the incidence estimates for age groups <1, 1-<2, 65-84, and ≥85 years old.

**Figure 3.**
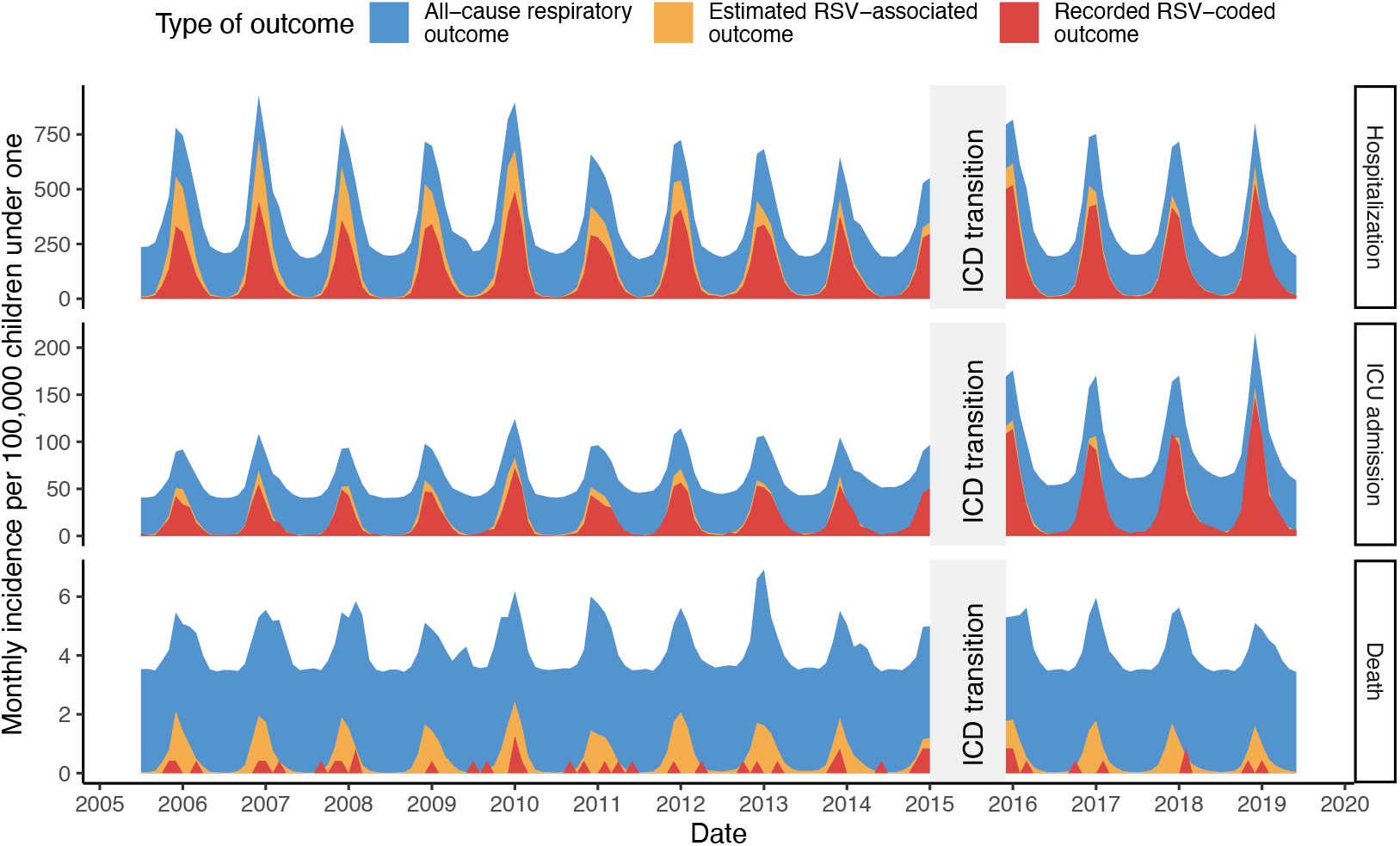
Monthly incidence of recorded RSV-coded, estimated RSV-associated, and estimated all-cause respiratory outcomes among infants under 1 year old, July 2005 - June 2019. The red area represents the incidence of outcomes recorded as being due to RSV in the HCUP database (RSV-coded outcomes). The yellow area represents the posterior median incidence of RSV-associated outcomes estimated from the model. The blue area represents the posterior median incidence of all-cause respiratory outcomes estimated from the model.

**Figure 4.**
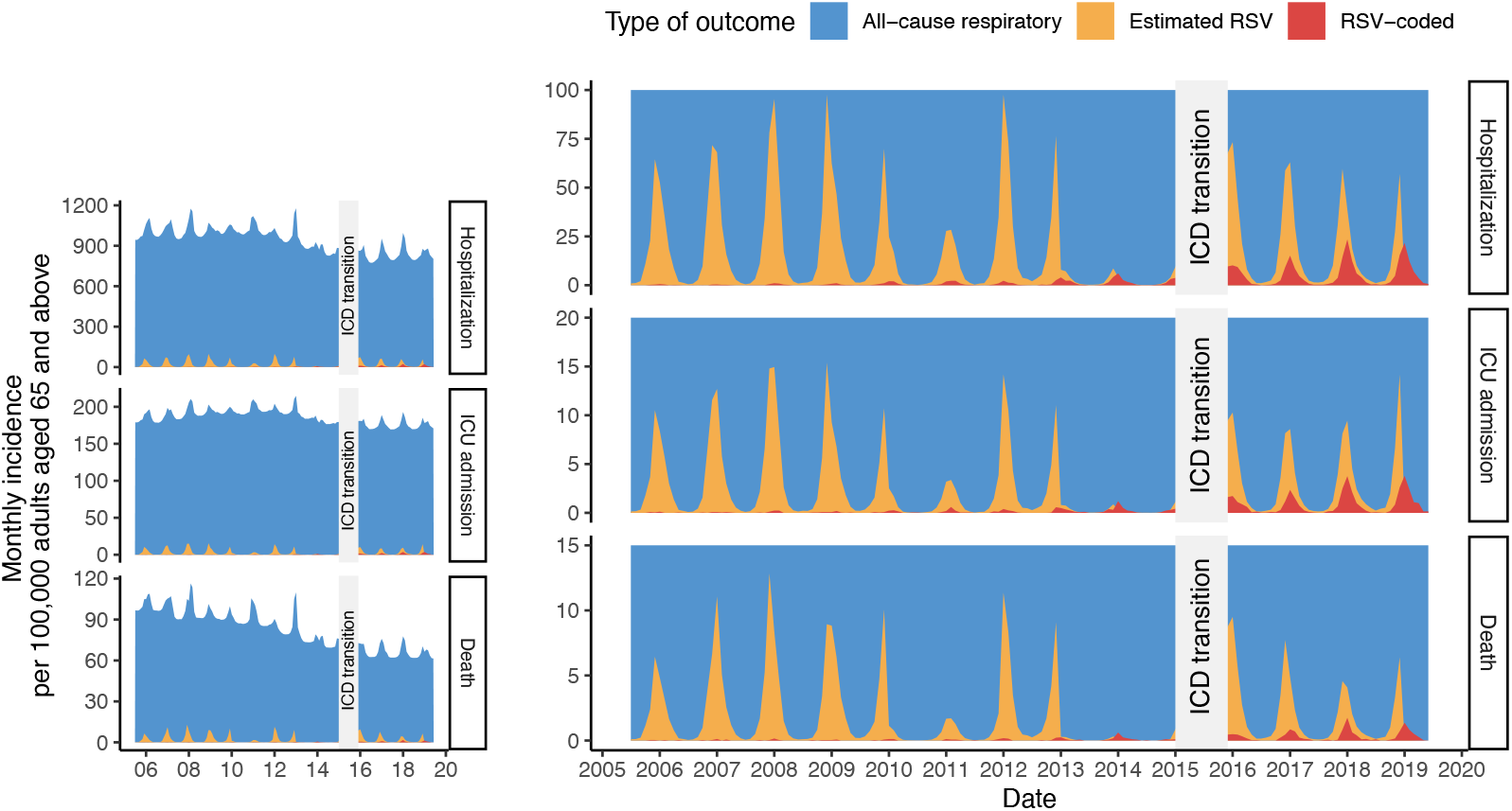
Monthly incidence of recorded RSV-coded, estimated RSV-associated, and estimated all-cause respiratory outcomes among adults aged 65 and above, July 2005 - June 2019 (left: original; right panel: zoomed in). The red area represents the incidence of outcomes recorded as being due to RSV in the HCUP database (RSV-coded outcomes). The yellow area represents the posterior median incidence of RSV-associated outcomes estimated from the model. The blue area represents the posterior median incidence of all-cause respiratory outcomes estimated from the model.

### Percent of respiratory outcomes attributable to RSV and influenza

RSV infections were estimated to contribute to a large proportion of respiratory hospitalizations, ICU admissions, and deaths in children <5 years old, especially in infants <1 year old, compared to influenza (Figure 5). Without stratification by SES, we estimated that 43.7% (95% CrI: 43.1-44.3%) of respiratory hospitalizations, 33.5% (95% CrI: 32.3-34.8%) of ICU admissions, and 13.1% (95% CrI: 10.7-15.8%) of deaths in the first year of life could be attributed to RSV infection. The attributable percent for all three outcomes was similar across SES groups. The attributable burden showed strong seasonality (Figure S10A). During the winter season, up to 60-80% of respiratory outcomes were estimated to be attributed to RSV among young children. For influenza, in contrast, the attributable percentage was low (<5%) for hospitalizations and ICU admissions. However, it was higher (10-20%) among school children (Figure 5). The attributable burden of influenza also exhibited seasonality, with a much higher attributable percentage during winter seasons (Figure S10B).

**Figure 5.**
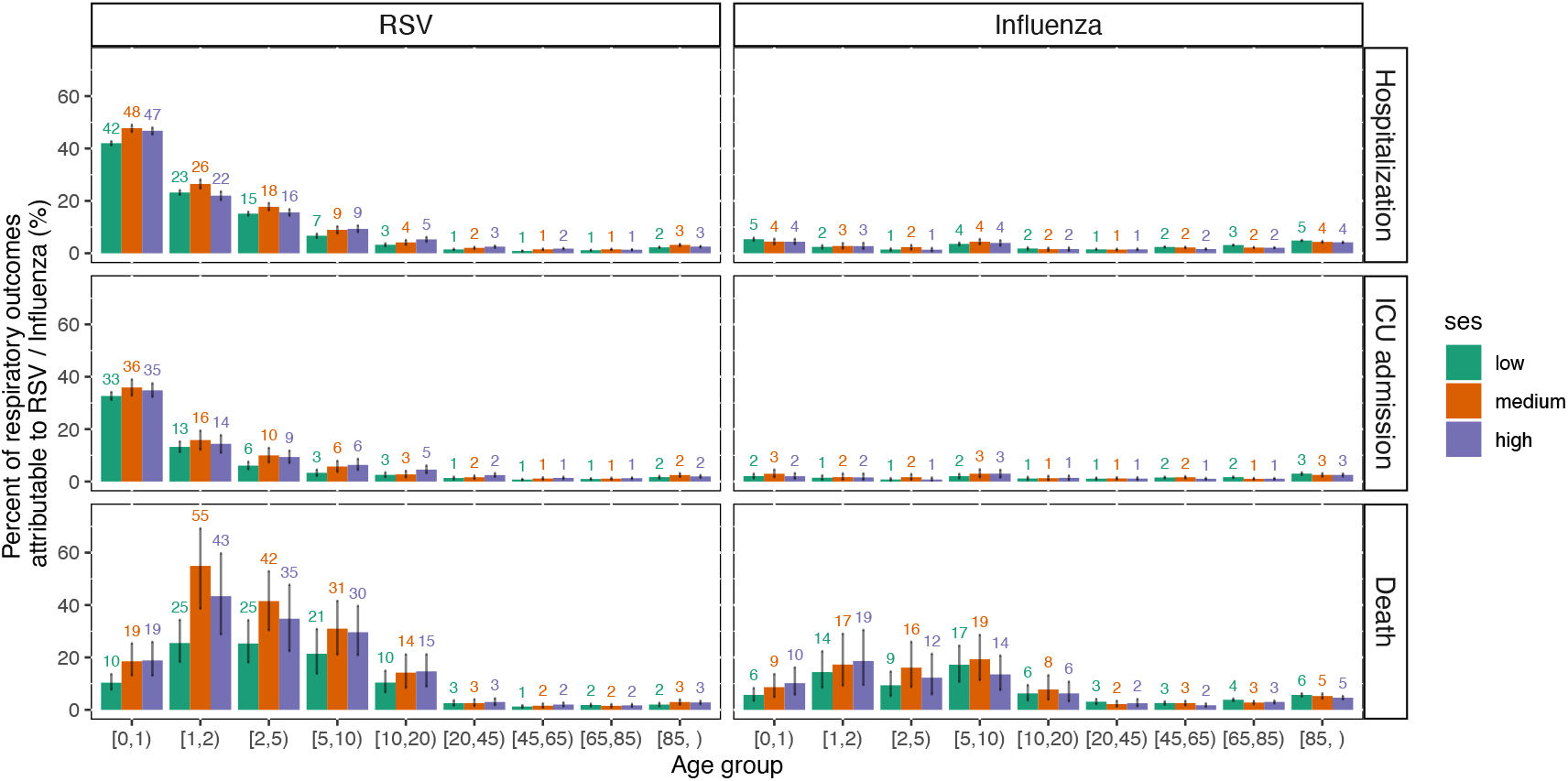
Percent of respiratory outcomes attributable to RSV and influenza infection, July 2005 - June 2019. The bars show the posterior median percent of all-cause respiratory outcomes (hospitalizations, ICU admissions, and deaths) that were estimated to be attributable to RSV or influenza infection, calculated as the estimated incidence of RSV-/influenza-associated outcomes divided by the estimated incidence of all-cause respiratory outcomes. The texts above the bars label the detailed numbers of the medians. The error bars indicate the 95% credible intervals of the estimated attributable percent.

### Inpatient ICU admission and mortality risk

The proportion of RSV- and influenza-associated hospitalizations resulting in ICU admission or death showed distinct patterns by age group (Figure 6). There was more variation by age in the ICU admission risk for RSV than for influenza (Figure S11A). The percent of RSV-associated hospitalizations admitted to the ICU was highest in the 65-84 year age group (17.5%), followed by the 45-64 year age group (15.6%) and infants <1 year (14.6%) (Figure S11A). The mortality risk of both RSV- and influenza-associated hospitalizations increased with age from less than 1% in the age groups <5 years to over 10% in the ≥65 years age group. We estimated a higher risk of ICU admission and death in the higher SES group for both RSV and influenza (Figure S11B), but this disparity persisted across all age groups only for the ICU admission risk of RSV (Figure 6A). The estimated proportion of hospitalizations admitted to the ICU was generally lower than the observed proportion directly calculated using the virus-coded outcomes for both RSV and influenza (Figure S12).

**Figure 6.**
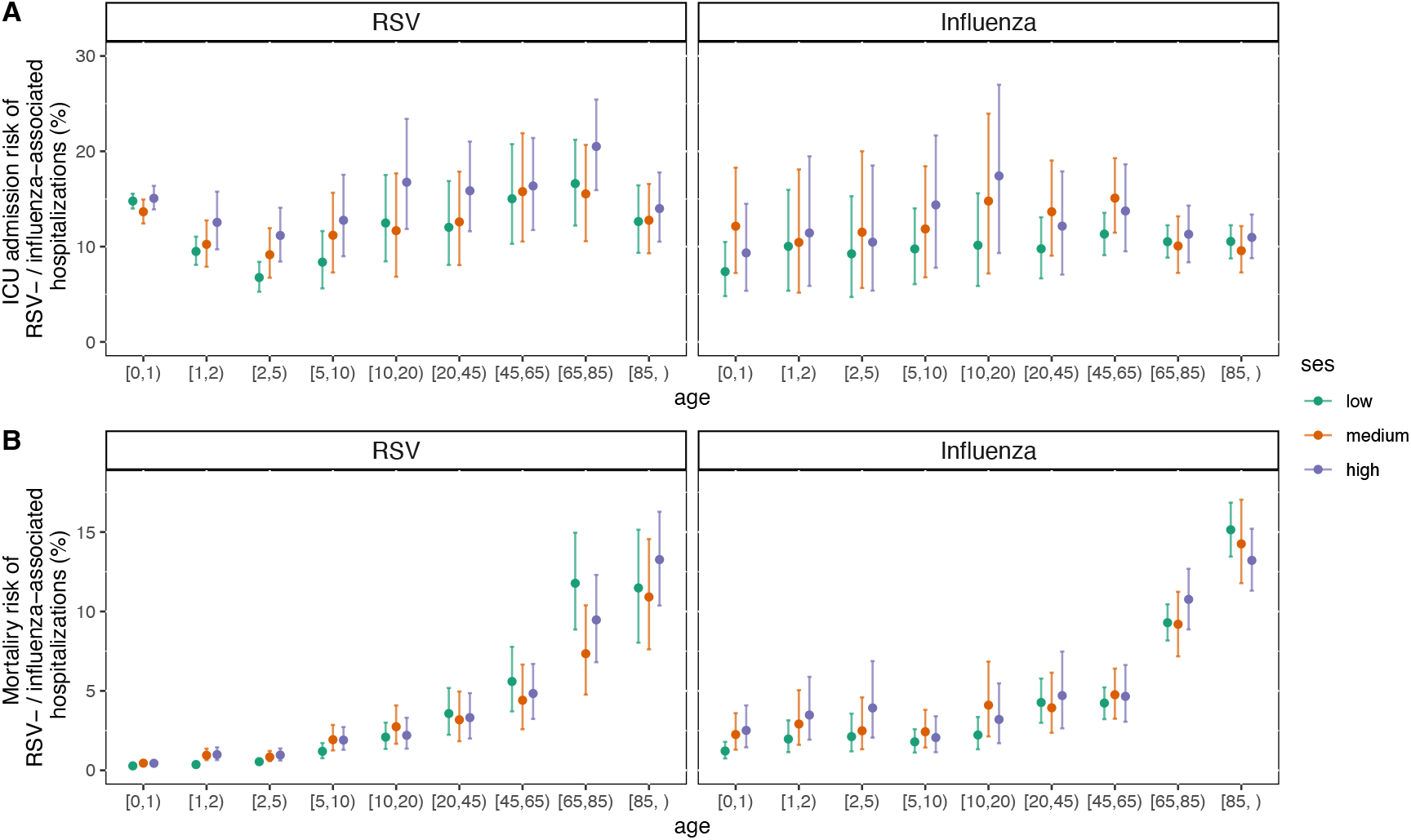
Estimated proportion of RSV- and influenza-associated hospitalizations admitted to the ICU admission (A) and resulting in death (B), July 2005 - June 2019. The dots indicate posterior median estimates of the proportion of RSV- and influenza-associated hospitalizations admitted to the ICU or dying. The error bars indicate the 95% credible intervals of the estimated ICU admission and mortality risk. ICU admission risk is defined as the ratio between the incidence of virus-associated ICU admission and the incidence of virus-associated hospitalizations; the mortality risk is defined as the ratio between the incidence of virus-associated deaths and the incidence of virus-associated hospitalizations.

### Recording fraction of RSV- and influenza-associated hospitalizations

Across all seasons, the percentage of estimated RSV-associated hospitalizations recorded as such was highest in infants <1 year (74.4%) and decreased with age, dropping to around 10% in adults (Figure 7A). However, over the study period, recording fractions rose in all age groups (Figure 7B). Among adults 65 years and older, the recording fraction increased from under 5% in the 2005/06 season to around 20% in the 2017/18 season and over 50% in the 2018/19 season.

**Figure 7.**
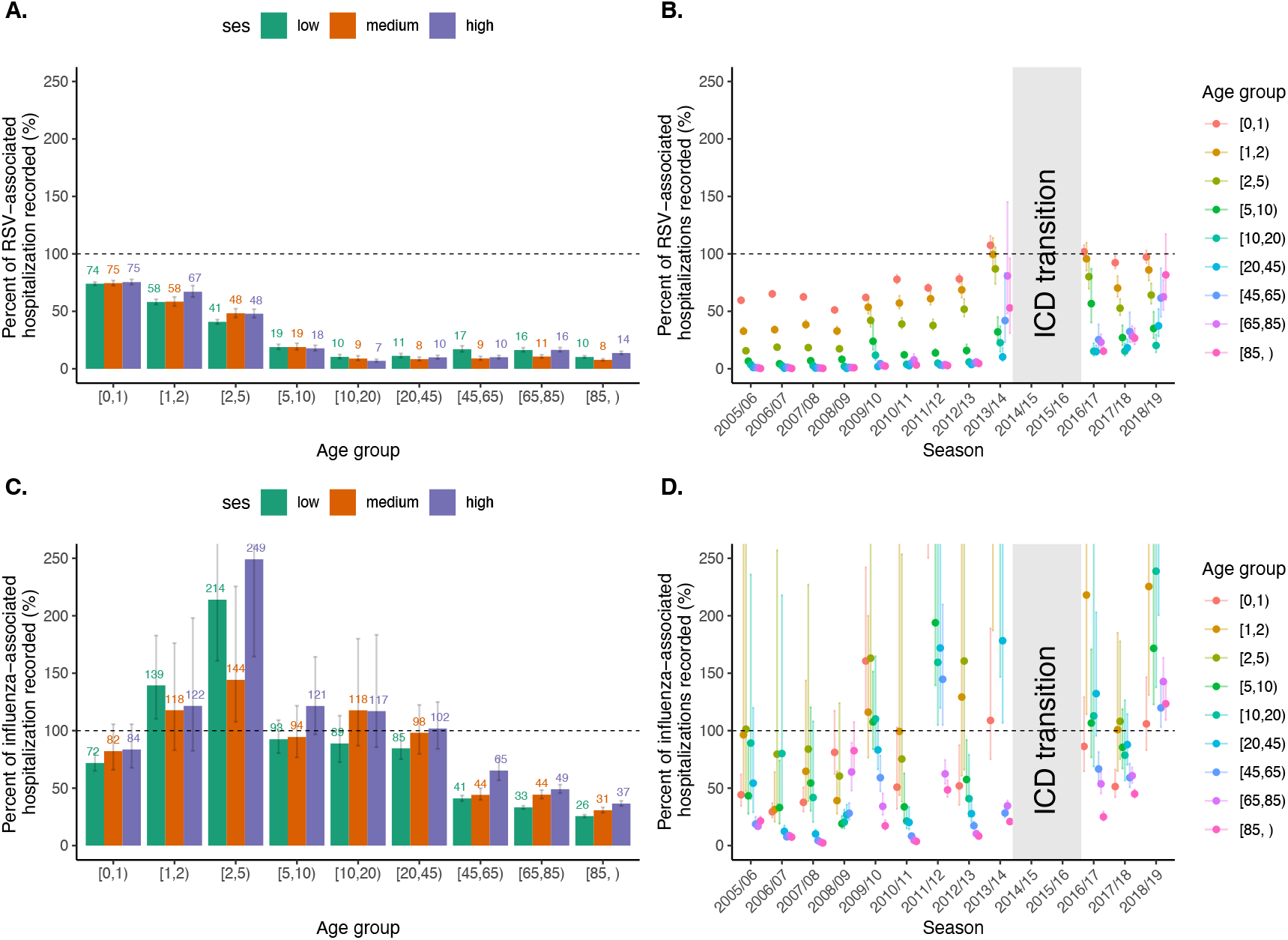
Percent of RSV-associated hospitalizations recorded as being due to RSV and influenza. Panel A) and C): Recording fraction of (A) RSV- and (C) influenza-associated hospitalizations, 2005-2019. The recording fraction is defined as the ratio of the number of RSV-coded and estimated RSV-associated hospitalizations. The bars represent the posterior median estimates of the recording fractions, and the error bars represent the 95% credible intervals of the estimates. Panel B) and D): Recording fraction of (B) RSV- and (D) influenza-associated hospitalizations over time by age. The dots represent the median estimates of the recording fraction, and the error bars represent 95% credible intervals of the estimates.

Our estimated recording fractions for influenza-associated hospitalizations exceeded 100% especially for children aged 2-4 years old (Figure 7C), possibly indicating an over-attribution of respiratory hospitalizations to influenza. Similar to RSV, the lowest recording fraction for influenza was also observed in the older age groups, although it was increasing over the study seasons (Figure 7D).

## Discussion

This study provided burden estimates of hospitalizations, ICU admissions, and deaths due to RSV and influenza by age and SES group. We also illustrated how the burden evolved over the study period from 2005 to 2019, which ended before RSV and influenza activities were disrupted by the COVID-19 pandemic and before the approval of several prophylactic products against RSV [22–25]. Our results demonstrated that 55% of RSV-associated hospitalizations and 61% of RSV-associated ICU admissions occurred in infants under 1, and 83% of RSV-associated deaths occurred in the ≥65 years age group. Older adults also had the highest influenza burden, with 72% of influenza-associated hospitalizations, 74% of influenza-associated ICU admissions, and 93% of influenza-associated deaths occurring in the ≥65-year age group. Although the recording fraction of RSV was found to be increasing for all ages over time, the gap between recorded and estimated RSV hospitalizations was still largest in older adults.

Our age-specific incidence estimates for RSV-associated hospitalizations in the US are generally consistent with previous studies [3,11,26–28]. Our estimates for older adults before the ICD transition in 2015 are slightly higher than previous estimates but are similar to those estimated using a model fit to the same database [11]. For seasons after 2015, our model provided comparable results to Branche et al., which also used data from New York and involved intensive clinical sampling among adults [27]. We reported similar estimates for death as in previous literature [3,29]. Our influenza-associated estimates across all three outcomes are also comparable with previous estimates in the literature [15,30–33], while offering greater granularity by providing estimates by SES and using finer age groups.

Assuming similar incidence across states, we extrapolate that 300,000 and 290,000 hospitalizations, 42,000 and 31,000 ICU admissions, 14,000 and 25,000 deaths due to RSV and influenza respectively would occur on average in the US every year. Among infants <1 year of age, 72,000 RSV-associated hospitalizations, 11,000 ICU admissions, and 240 deaths would occur annually. Among adults aged 65 and above, 97,000 RSV-associated hospitalizations, 16,000 ICU admissions, and 10,000 deaths would occur annually. Additionally, 190,000 influenza-associated hospitalizations, 19,000 ICU admissions, and 21,000 deaths would occur in adults aged 65 and older.

Our study examined RSV and influenza burden across different SES groups classified using supervised PCA that incorporated several socioeconomic variables. A previous study by Zheng et al. classified SES by household income and reported a higher incidence of RSV-associated hospitalizations in the low-income group among all age groups. We found the same trend by SES in the incidence of RSV hospitalizations and ICU admissions only among children under 10, but not in older age groups, and we did not find a consistent pattern of influenza incidence by SES. There have been mixed findings on the impact of SES on respiratory disease burden among adults [5,34,35]. One study conducted in New Zealand found that living in low SES neighborhoods was associated with increased RSV hospitalization rates in adults [36]. Other studies found SES was associated with increased incidence of RSV- and influenza-associated hospitalizations, but not with more severe outcomes such as ICU admission and death [34,37]. Our model also estimated a higher ICU admission risk and mortality risk in the higher SES group for both pathogens, and this disparity persisted in all age groups in ICU admission risk for RSV (Figure 6). The ICU admission risk directly calculated from the RSV-coded outcomes showed a similar trend (Figure S12B). This might be due to the disparity in the ICU capacity between hospitals and the impact of a family’s SES on the decision to admit patients into ICUs. Another possible reason is that the threshold for admission to hospital might be lower for low-SES individuals, and this would inflate the denominator for ICU admission risk.

Our results showed that the proportion of RSV-associated hospitalizations admitted to the ICU had a larger variation by age for RSV, compared to a rather stable trend by age for influenza. Relatively constant ICU admission risks of influenza-associated hospitalizations by age were also estimated by O’Halloran et al.[33]. Previous studies on RSV disease progression have predominantly focused on in-hospital ICU admission and mortality risks, relying on individual-level clinical data. Walsh et al. reported an ICU admission risk of 28% among adults hospitalized due to RSV (2014-2016) and did not observe a significant difference by age [38]. A recent study among US adults reported that 24.8% of RSV hospitalizations were admitted to the ICU and 12.0% required IMV or died [13], which is higher than our direct estimates of 17.2% ICU admission and 5.0% mortality risk among adults calculated directly from the virus-coded outcomes (Figure S12), and higher still than our estimates from the model that accounted for underreporting. This indicates that our model additionally captured less severe hospitalizations that were missed by healthcare facilities or incorrectly coded as due to other causes.

We highlighted an elevated incidence of RSV- and influenza-associated outcomes among adults 65 years and older for both RSV and influenza and found that the inpatient mortality risk was also considerably higher for both pathogens in this age group. Our results again captured the significantly lower recording fraction of RSV hospitalizations in this age group as indicated by Zheng et al.[11]. Additionally, we observed an increasing trend of recording fractions of RSV in all ages over the study period, which has also been reported in other studies [11,39]. This might be correlated with the changes in testing practices over the study period.

Our incidence estimates over time showed that RSV-associated ICU admissions in children under 2 years old almost doubled over the studied period, despite a moderate decrease in RSV-associated hospitalizations. A similar trend was also observed in the Netherlands, which reported a fourfold increase in pediatric ICU admissions for RSV bronchiolitis among children under 2 years old from 2003 to 2016 [40]. We found that this increase was accompanied by a decreasing length of in-hospital stay for ICU-admitted RSV cases, an increasing number receiving NIMV, and a decreasing number receiving IMV over the years in this age group (Figure S7, S8). Changes in the options of ventilation support upon ICU admissions might have influenced the ICU admission threshold, leading to less severe cases being admitted to the ICU over more recent seasons. What the admission criteria are, and how they are evolving in clinical practices, is often complicated and requires further study with more detailed data from clinical settings.

### Limitations

Our study has several limitations. First, the time series data we used included a one-year gap in 2015 due to the transition from ICD-9 to ICD-10. To avoid interference between the pre-and post-transition periods, we did not incorporate any auto-regressive structure by season in the model. Although the transition to ICD-10 introduced an additional category for acute bronchitis due to RSV (J20.5), this category was previously encompassed within one of the three ICD-9 codes for RSV and thus is not expected to significantly impact case classification. Nonetheless, caution is warranted when comparing estimates from these two periods. Additionally, the diagnosis of hospitalizations based solely on ICD codes is susceptible to misclassification, particularly since RSV and influenza share similar symptoms with many other respiratory infections.

Second, the classification of ICU admission was not based on clinical records but the ICU-related revenue codes in the medical charge dataset, as they were the only available indicators for ICU admission in the database. Some patients might only stay in the ICU briefly and later be transferred out but were also counted as admitted to the ICU. The gold standard defines ICU admission only if a patient remains in the ICU for at least 2 consecutive hours or longer [17]. Although studies have shown revenue codes provide high sensitivity and specificity in identifying true ICU admissions, our classification might additionally capture some less severe cases.

Third, our finding of the increasing recording fraction relied on the validity of estimates of RSV-associated hospitalizations, which might be hard to validate. Nevertheless, our estimates of hospitalizations generally aligned with previous studies [11]. Furthermore, we not only observed an increasing recording fraction in all age groups, including the youngest age group for which estimates were usually more reliable due to more frequent testing. Our model also estimated that the recording fraction of influenza for children aged 2-4 years old was larger than 100%, indicating that respiratory hospitalizations in this age group might have been overly attributed to influenza, thus leading to underestimation of RSV incidence in this age group.

## Conclusions

This study provided a comprehensive overview of the inpatient burden of RSV and influenza before the COVID-19 pandemic. We found that RSV and influenza disproportionately affected different age groups and exhibited varying impacts across levels of disease severity. Our results showed that infants under 1 year of age had the highest incidence of RSV-associated hospitalizations and ICU admissions, while older adults aged 65 and above had the highest incidence of RSV-associated death. Older adults additionally experienced a higher incidence of influenza-associated hospitalizations, ICU admissions, and deaths. Although the largest gap between recorded and estimated RSV hospitalizations was observed in older adults, the recording fraction for all age groups has been increasing over the study period. The proportion of RSV- and influenza-associated hospitalizations admitted to the ICU varied by age, and the mortality risk was considerably higher in older adults.

## Data Availability

Data produced in the present study are available upon reasonable request to the authors.

## Acknowledgments

Thanks to Lone Simonsen for helpful feedback on these analyses.

