## Supplementary Materials for "Estimating the burden of RSV- and influenza-associated hospitalizations, ICU admissions, and deaths across age and socioeconomic groups in New York State, 2005-2019"

### Supplementary Methods

#### Hierarchical Bayesian regression model

We used hierarchical Bayesian regression models to estimate the incidence of different metrics measuring RSV- and influenza-associated clinical burden: hospitalizations, ICU admissions and deaths, by age and SES group. The estimated mean count of all-cause respiratory hospitalizations, ICU admissions and deaths in age group $j$ of SES group $k$ in month $i$, $\lambda_{ijk}^{hosp}$, $\lambda_{ijk}^{ICU}$, and $\lambda_{ijk}^{death}$ were given as

$$\lambda_{ijk}^{hosp}/\,pop_{ijk} = \beta_{0jk}^{hosp} + \alpha_{1y(i)jk}^{hosp} + \alpha_{2m(i)}^{hosp} + \beta_{1y(i)jk}^{hosp}RSV_{ik}/\,pop_{ijk} + \beta_{2y(i)jk}^{hosp}Flu_{ik}/\,pop_{ijk}$$

$$\lambda_{ijk}^{ICU}/\,pop_{ijk} = \beta_{0jk}^{ICU} + \alpha_{1y(i)jk}^{ICU} + \alpha_{2m(i)}^{ICU} + \beta_{1y(i)jk}^{ICU}RSV_{ik}/\,pop_{ijk} + \beta_{2y(i)jk}^{ICU}Flu_{ik}/\,pop_{ijk}$$

$$\lambda_{ijk}^{death}/\,pop_{ijk} = \beta_{0jk}^{death} + \alpha_{1y(i)jk}^{death} + \alpha_{2m(i)}^{death} + \beta_{1y(i)jk}^{death}RSV_{ik}/\,pop_{ijk} + \beta_{2y(i)jk}^{death}Flu_{ik}/\,pop_{ijk}$$

in which $\beta_{0jk}$ is the underlying intercept parameter, $\alpha_{1y(i)jk}$ represents the underlying temporal trend varying over the epidemic seasons $y(i)$, and $\alpha_{2m(i)}$ represents the monthly variation in each month $m(i)$, capturing seasonality. We assume the temporal trends $\alpha_{1y(i)jk}$ varies by age group $j$ of SES group $k$, and seasonality $\alpha_{2m(i)}$ is shared by all ages and SES groups.

$RSV_{ik}$ is the indicator for RSV, defined as the number of RSV-coded hospitalizations in age groups under 2 years old in month $i$ and SES group $k$, and $Flu_{ik}$ is the indicator for influenza, defined as the number of influenza hospitalizations in all ages in month $i$ and SES group $k$. $\beta_{1y(i)jk}$ and $\beta_{2y(i)jk}$ are the coefficients for $RSV_{ik}$ and $Flu_{ik}$, respectively. These effects vary by epidemic season, $y(i)$, allowing for the association between the viral indicators and the outcome to change over time and between seasons due to differences in testing and differences in severity of the dominant strains. By multiplying the scaling factors with the incidence of virus-associated hospitalizations, we get $\beta_{1y(i)jk}RSV_{ik}$ and $\beta_{2y(i)jk}Flu_{ik}$, which represent the estimated incidence of RSV- and influenza-associated disease outcomes (hospitalizations, ICU admissions, deaths), respectively. The incidence terms are adjusted for the population size $pop_{ijk}$ . We assume the observed all-cause respiratory hospitalizations: $Y_{ijk}^{hosp}$, ICU admissions: $Y_{ijk}^{ICU}$ , and deaths: $Y_{ijk}^{death}$ follow Poisson distributions such that:

$$Y_{ijk}^{hosp} \sim Poisson(\lambda_{ijk}^{hosp})$$

$$Y_{ijk}^{ICU} \sim Poisson(\lambda_{ijk}^{ICU})$$

$$Y_{ijk}^{death} \sim Poisson(\lambda_{ijk}^{death})$$

To stabilize the model performance, the three clinical outcomes were linked together through RSV- and influenza-associated disease progression risks, taking the rationale that in-hospital RSV-associated ICU admissions and deaths are subsets of RSV-associated hospitalizations (and likewise for influenza). The relationships thus can be given as:

$$\beta_{1y(i)jk}^{ICU} = \beta_{1y(i)jk}^{hosp} * p_{1y(i)jk}^{ICU}$$

$$\beta_{1y(i)jk}^{death} = \beta_{1y(i)jk}^{hosp} * p_{1y(i)jk}^{death}$$

$$\beta_{2y(i)jk}^{ICU} = \beta_{2y(i)jk}^{hosp} * p_{2y(i)jk}^{ICU}$$

$$\beta_{2y(i)jk}^{death} = \beta_{2y(i)jk}^{hosp} * p_{2y(i)jk}^{death}$$

in which $p_{1y(i)jk}^{ICU}$ and $p_{1y(i)jk}^{death}$ represent the proportion of RSV-associated hospitalizations resulting in ICU admission and death, respectively, with variation by epidemic season $y(i)$, age group $j$, and SES group $k$. Similarly, $p_{2y(i)jk}^{ICU}$ and $p_{2y(i)jk}^{death}$ represent the ICU admission risk and mortality risk of influenza-associated hospitalizations.

We ensured positivity for the intercept parameters ($\beta_0$) for the two pathogens:

$$\beta_{0jk} = exp\ \{\mu_{0jk}\}$$

$$\mu_{0jk} \sim N(\mu_0, \sigma_0^2)$$

We also ensured positivity for the temporal variation term $\alpha_{1y(i)jk}$ and seasonal variation term $\alpha_{2m(i)}$, such that

$$\alpha_{1y(i)jk} = exp\ \{\mu_{1y(i)jk}\}$$

$$\mu_{1y(i)jk} \sim N(0, \sigma_1^2)$$

$$\alpha_{2m(i)} = exp\ \{\mu_{2m(i)}\}$$

$$\mu_{2m(i)} \sim N(0, \sigma_2^2)$$

For the coefficients of RSV- and influenza-coded hospitalizations, $\beta_1$ and $\beta_2$, we modeled them as a multiplicative combination of seasonal, age, and SES effects, such that

$$\beta_{1y(i)jk} = exp\ \{\varphi_{1y(i)} + \omega_{1j} + \gamma_{1k} + \epsilon_{1y(i)jk}\}$$

$$\epsilon_{1y(i)jk} \sim N(0, \sigma_{\epsilon_1}^2)$$

$$\beta_{2y(i)jk} = exp\ \{\varphi_{2y(i)} + \omega_{2j} + \gamma_{2k} + \epsilon_{2y(i)jk}\}$$

$$\epsilon_{2y(i)jk} \sim N(0, \sigma_{\epsilon_2}^2)$$

in which $\omega_{1j}$ and $\omega_{2j}$ represent the age group effects, $\gamma_{1k}$ and $\gamma_{2k}$ represent the SES effects, $\varphi_{1y(i)}$ and $\varphi_{2y(i)}$ represent the epidemiologic year effects potentially due to differences in the circulating viral strain or subtypes, and $\epsilon_{1y(i)jk}$ and $\epsilon_{2y(i)jk}$ represent unexplained variation.

We modeled the group-specific ICU admission risk, $p_{y(i)jk}^{ICU}$ , and mortality risk, $p_{y(i)jk}^{death}$, to be centered around shared Beta distributions, such that

$$p_{y(i)jk}^{ICU} \sim Beta(1, \theta^{ICU})$$

$$p_{y(i)jk}^{death} \sim Beta(1, \theta^{death})$$

Other hyperparameters were weakly informative as follows:

$$\omega_{1j} \sim N(0, \sigma_{\omega_1}^2)$$

$$\gamma_{1k} \sim N(0, \sigma_{\gamma_1}^2)$$

$$\varphi_{1y(i)} \sim N(0, \sigma_{\varphi_1}^2)$$

$$\omega_{2j} \sim N(0, \sigma_{\omega_2}^2)$$

$$\gamma_{2k} \sim N(0, \sigma_{\gamma_2}^2)$$

$$\varphi_{2y(i)} \sim N(0, \sigma_{\varphi_2}^2)$$

$$\sigma_0^2, \sigma_1^2, \sigma_2^2, \sigma_{\omega_1}^2, \sigma_{\omega_2}^2, \sigma_{\gamma_1}^2, \sigma_{\gamma_2}^2, \sigma_{\varphi_1}^2, \sigma_{\varphi_2}^2, \sigma_{\epsilon_1}^2, \sigma_{\epsilon_2}^2, \theta^{ICU}, \theta^{death} \sim Inverse\ Gamma(0.01, 0.01)$$

The model was fitted using `rjags` package in R Software, version 4.3.1[2]. Posterior samples were collected using a Markov chain Monte Carlo (MCMC) algorithm. To make posterior inference, 10,000 MCMC iterations were collected following a burn-in period of 60,000 iterations of each chain. The combined interactions from three chains were used to calculate burden estimates. Convergence was assessed using trace plots[1]. Uncertainty was quantified using 95% credible intervals.

### Summarizing model outputs

The incidence of specific outcomes (hospitalizations, ICU admissions, deaths) associated with RSV and influenza infection was calculated by multiplying each virus-coded outcome by its estimated coefficient (i.e. scaling factor) and then dividing by the population size of the specific risk group, i.e. age and SES group. The percent of the all-cause respiratory outcomes that could be attributed to RSV and influenza infection was calculated as the estimated number of virus-associated outcomes divided by the estimated number of all-cause respiratory cases of the specific risk group. We defined the recording fraction of virus-associated hospitalizations as the percent of estimated virus-associated hospitalizations recorded as being due to the specific virus. This was calculated as the ratio of the number of virus-coded hospitalizations recorded in the HCUP database and the estimated number of virus-associated hospitalizations from the model. Disease progression risks (i.e., ICU admission risk, defined as the ratio of ICU admissions and hospitalizations, and mortality risk, calculated as the ratio of deaths and hospitalizations) were also estimated from the model.

### Categorize SES using Principal Component Analysis (PCA)

We used the socioeconomic status (SES) of the ZIP code where the patients resided as an approximate measure for the SES status of that patient due to the lack of individual-level socioeconomic data in the State Inpatient Databases of the Healthcare Cost and Utilization Project (HCUP) dataset. We extracted data on the employment rate, average household size, median household income level, population density, proportion of population that were Black or African American, and proportion of population below the poverty line for each ZIP code in New York state from the US Census Bureau's American Community Survey [3]. Supervised PCA was performed using the incidence of RSV hospitalization at the ZIP-code level as the target variable. We classified all the ZIP codes into three groups (low, medium, and high) utilizing the first PCA component, which accounted for the majority (~50%) of the variance. The scree plot and loading plot of the PCA are shown in Figure S1. Distributions of included variables by classified SES are shown in Figure S2. The distribution of ZIP codes by classified SES group is shown in Figure S3.

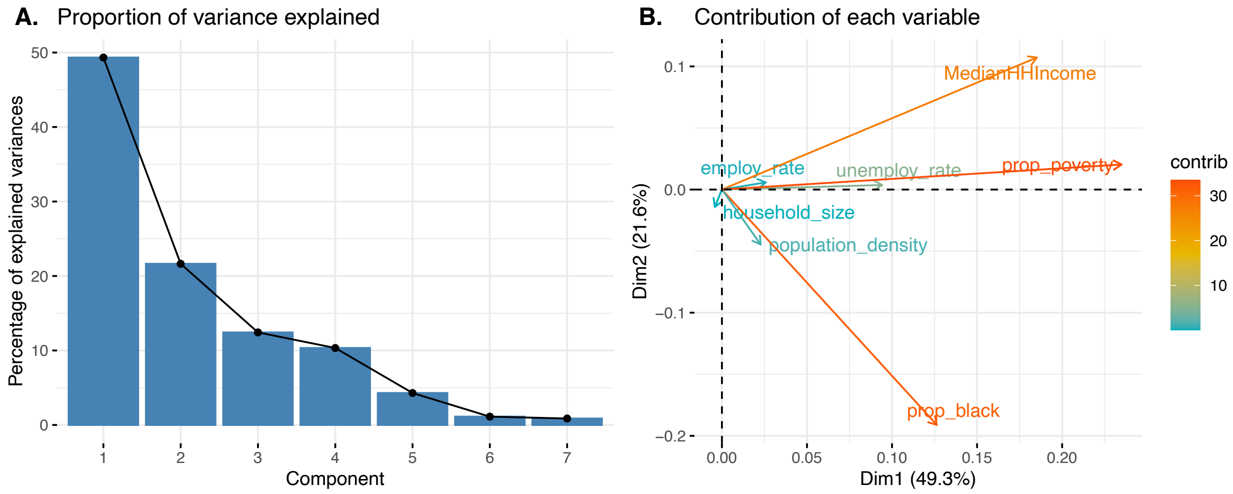


**Figure S1. Scree plot (A) and loading plot (B) of the supervised PCA classifying the SES group of the patients' residential ZIP codes.** The scree plot (A) shows the proportion of variance explained by each of the PCA components. The loading plot (B) shows the contribution of each variable included in the supervised PCA (MedianHHIncome: median household income; prop_poverty: proportion of the population living under the poverty line; prop_black: proportion of the population that are Black or African American; employ_rate: proportion of population that are employed; unemploy_rate: proportion of the population that are unemployed; household_size: average number of people living in each household; population density: persons per square mile). The length of the variable vector represents the strength of the variable's contribution to the principal components, which is also shown by the colors of the vectors.

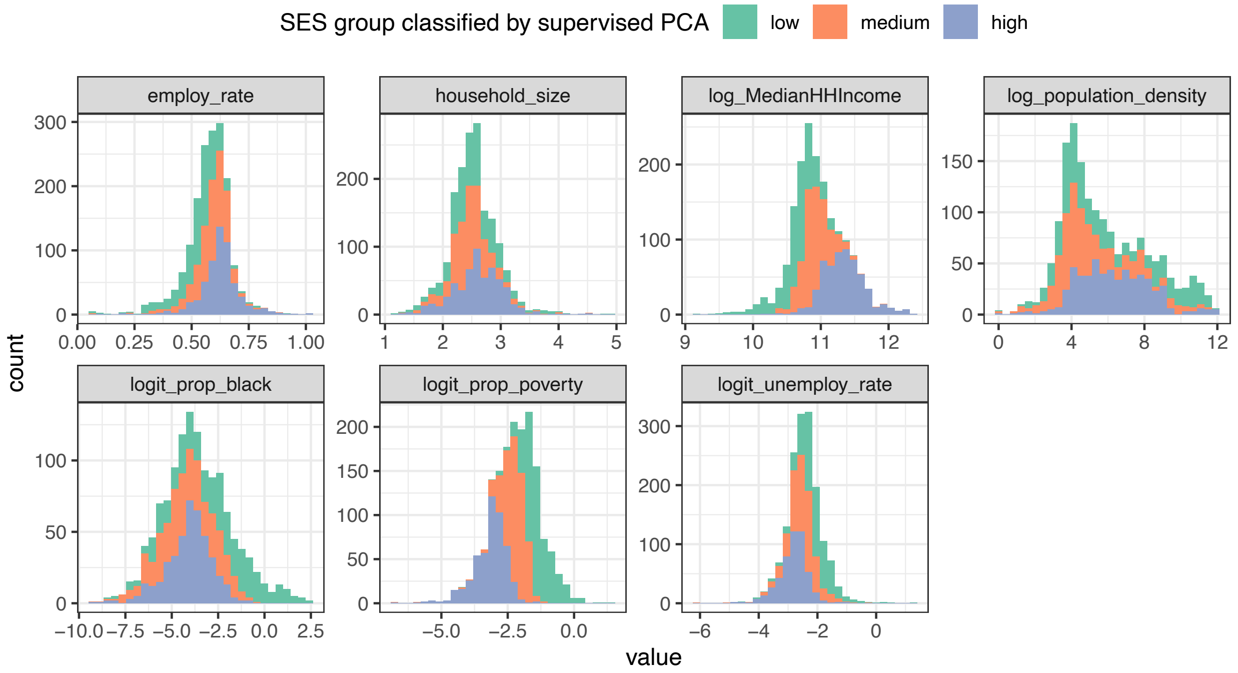


**Figure S2. Distribution of variables included in the supervised PCA classifying SES group of patients' residential zip codes.** Each panel plots the distribution of an included variable, with the colors of the bars representing the three SES groups classified by PCA. MedianHHIncome: median household income; prop_poverty: proportion of the population living under the poverty line; prop_black: proportion of the population that is Black or African American; employ_rate: proportion of the population that are employed; unemploy_rate: proportion of the population that are unemployed; household_size: average number of people living in each household; population density: persons per square mile. Some variables were rescaled using log or logit transformation due to the skewness of the data.

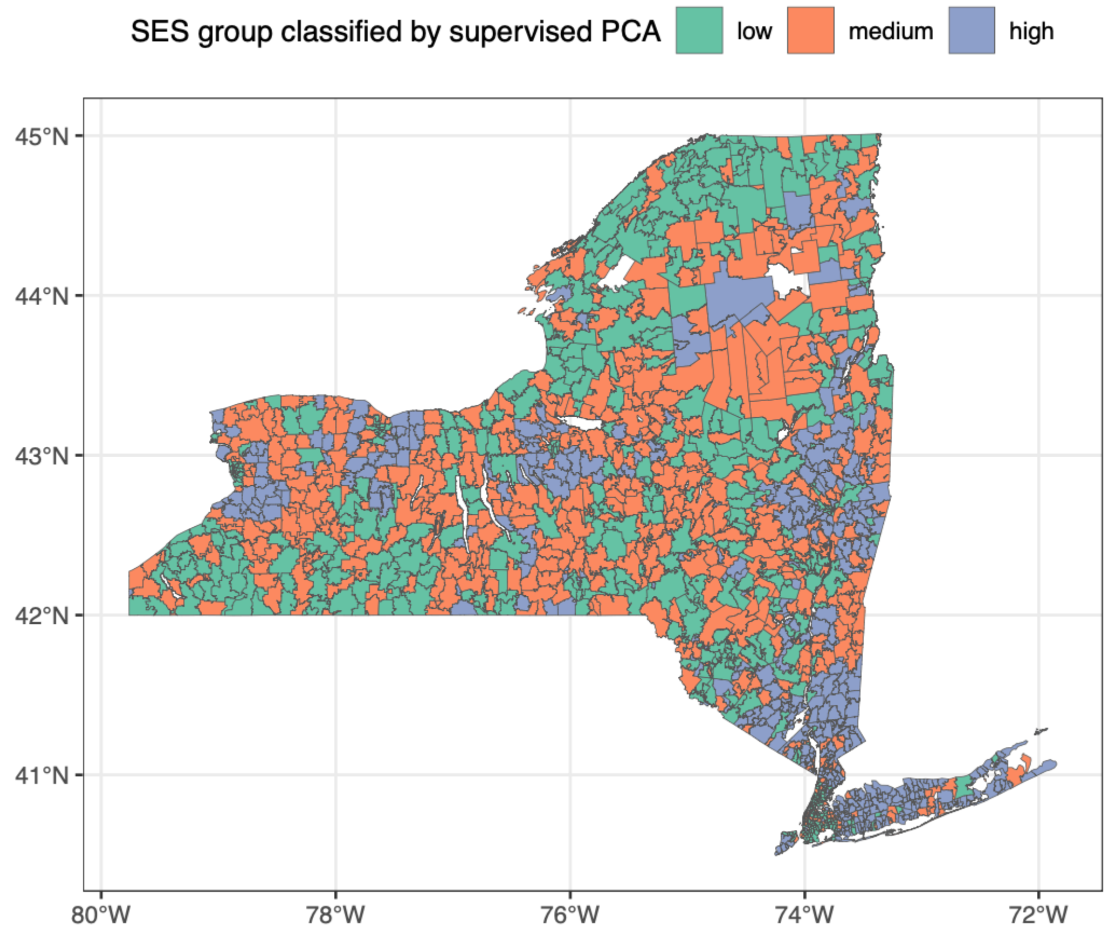

**Figure S3. Distribution of ZIP codes in New York state by PCA-classified SES group.**

### Supplementary Results

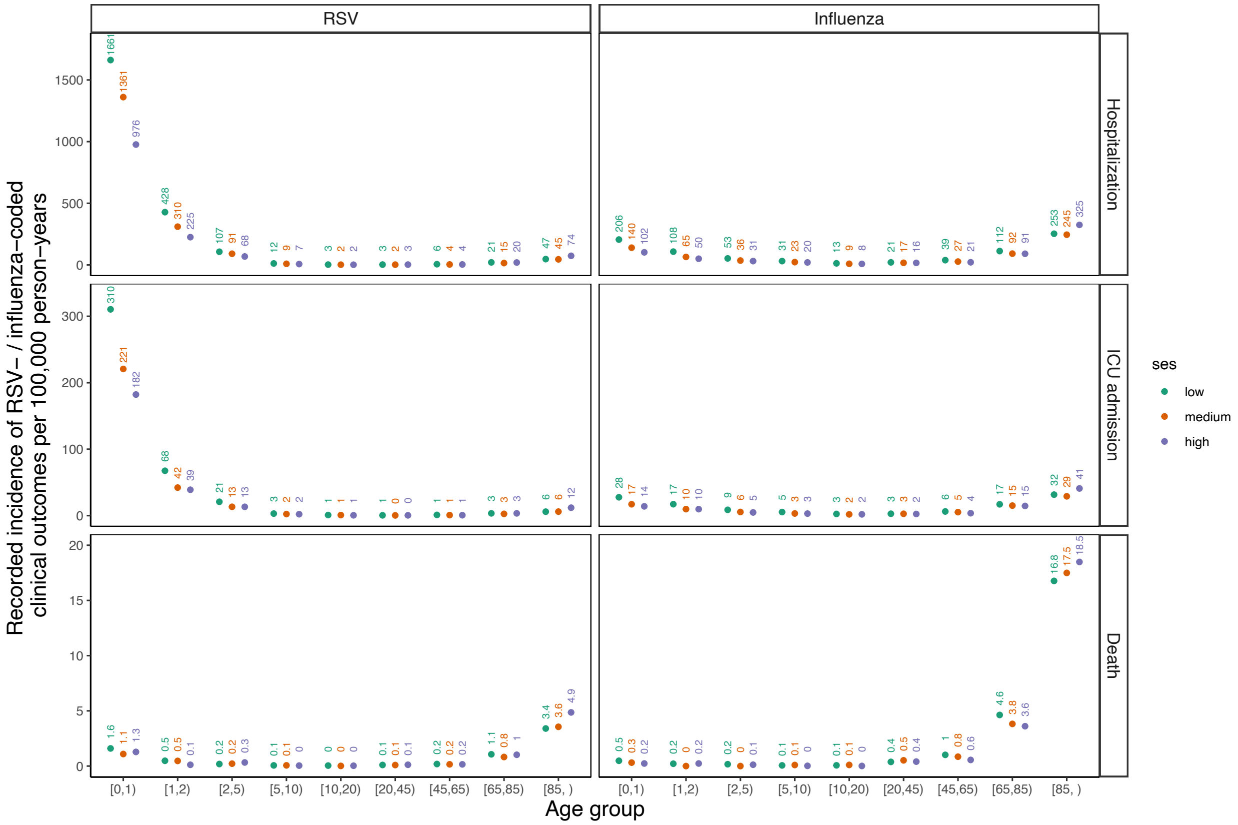


**Figure S4. Annual incidence of RSV- and influenza-coded outcomes recorded in the Healthcare Cost and Utilization Project (HCUP) dataset, 2005-2019.** The incidence of virus-coded outcomes here was directly calculated using the annual number of outcomes (i.e., hospitalizations, ICU admissions, deaths) that were coded (by ICD codes) as being due to a specific virus in a specific risk group, divided by the population size of that risk group.

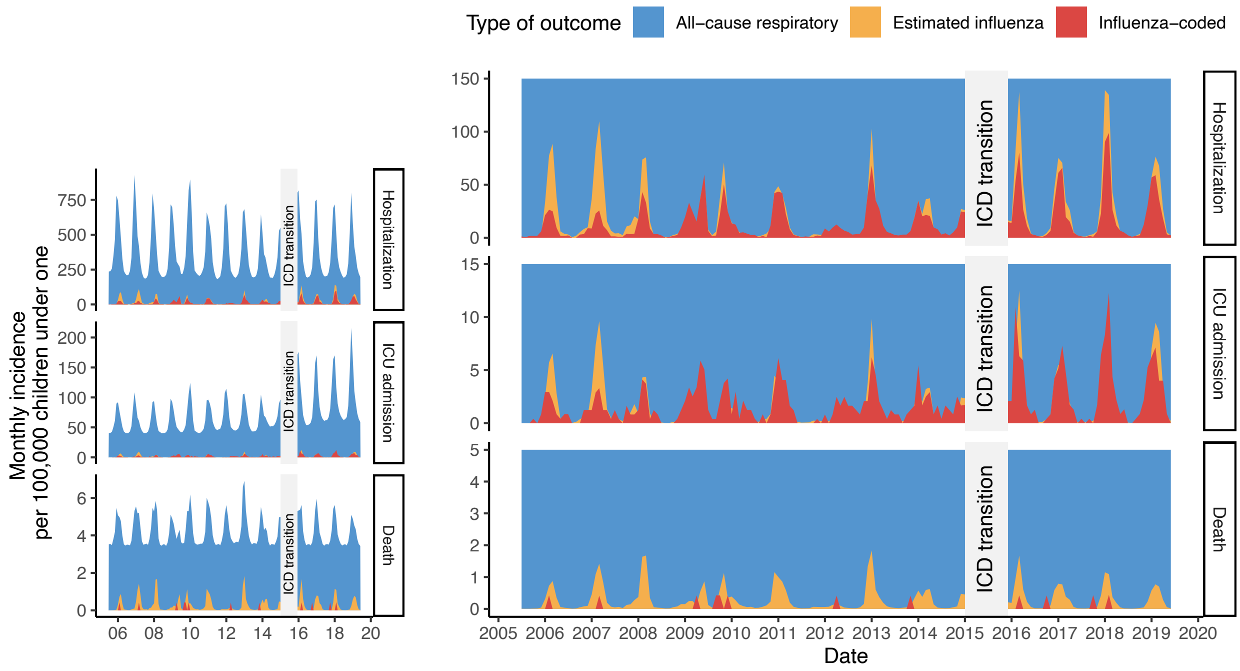


**Figure S5. Monthly incidence of recorded influenza-coded, estimated influenza-associated, and estimated all-cause respiratory outcomes among infants under 1 year old, July 2005 - June 2019 (left: original; right: zoomed in).** The red area represents the incidence of outcomes recorded as being due to influenza in the HCUP database (influenza-coded outcomes). The yellow area represents the median incidence of influenza-associated outcomes estimated from the model. The blue area represents the median incidence of all-cause respiratory outcomes estimated from the model.

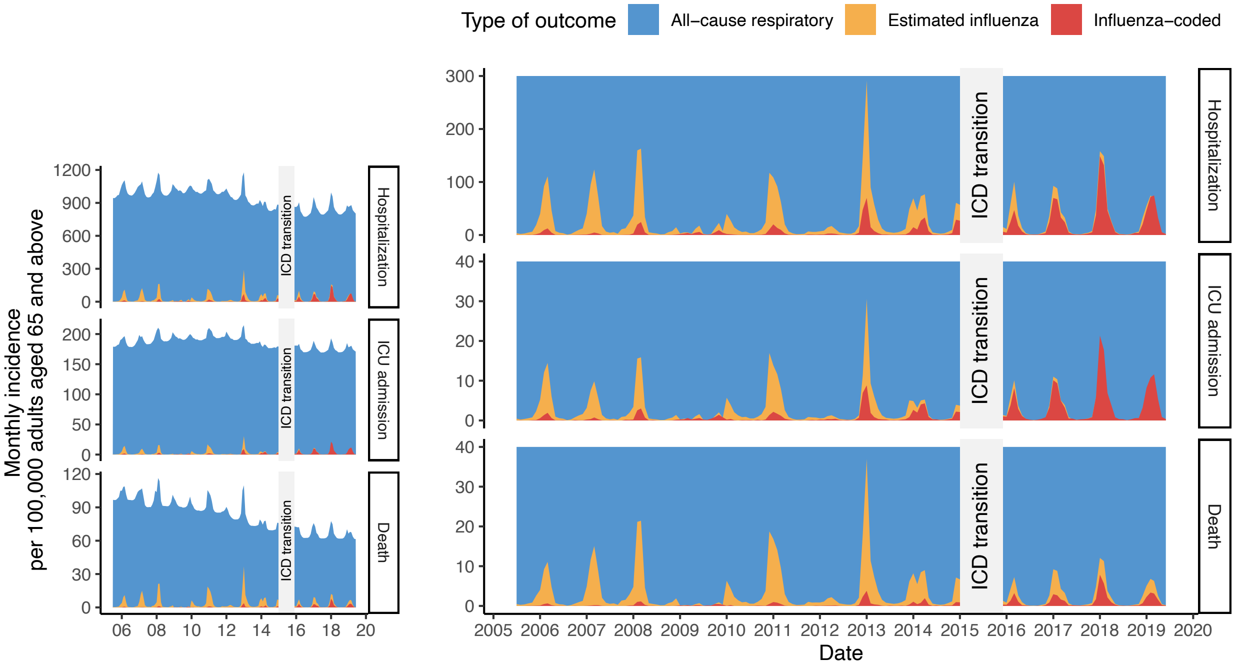


**Figure S6. Monthly incidence of recorded influenza-coded, estimated influenza-associated, and estimated all-cause respiratory outcomes among adults aged 65 and older, July 2005 - June 2019 (left: original; right: zoomed in).** The red area represents the incidence of outcomes recorded as being due to influenza in the HCUP database (influenza-coded outcomes). The yellow area represents the median incidence of influenza-associated outcomes estimated from the model. The blue area represents the median incidence of all-cause respiratory outcomes estimated from the model.

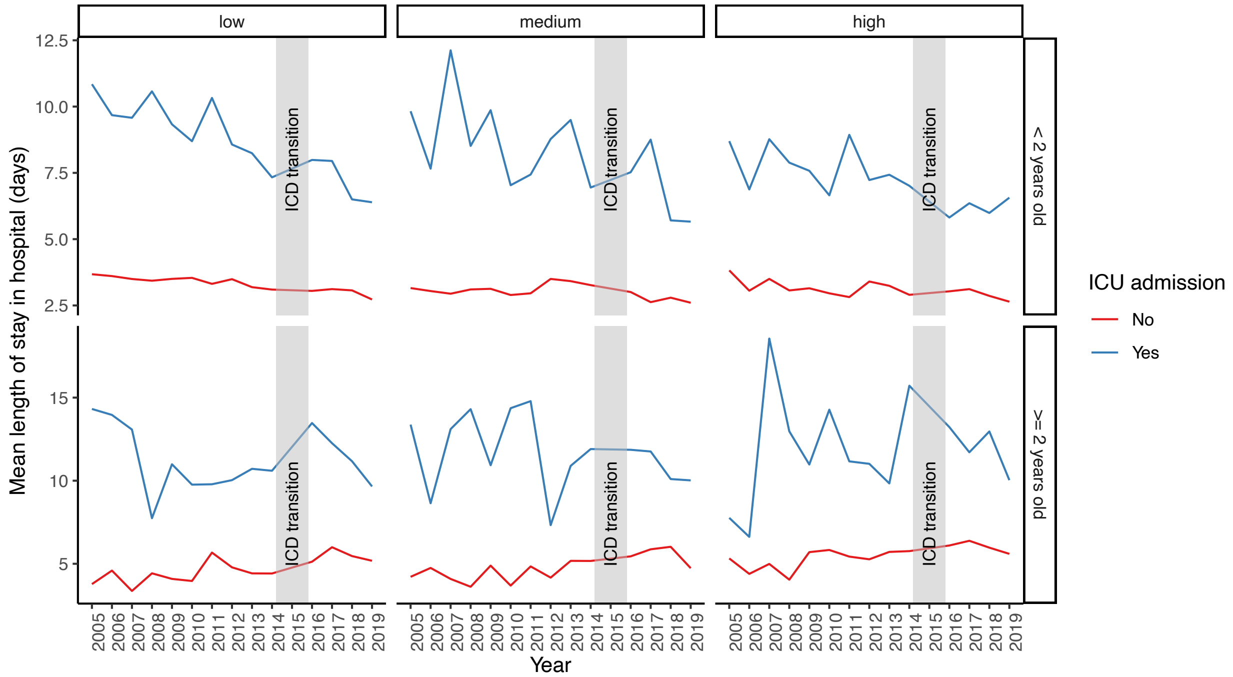


**Figure S7. The mean length of stay in hospital (in days) of RSV-associated hospitalizations over time, by socioeconomic status (SES).** The three vertical panels show the hospital length of stay among patients in low, medium, and high SES groups.

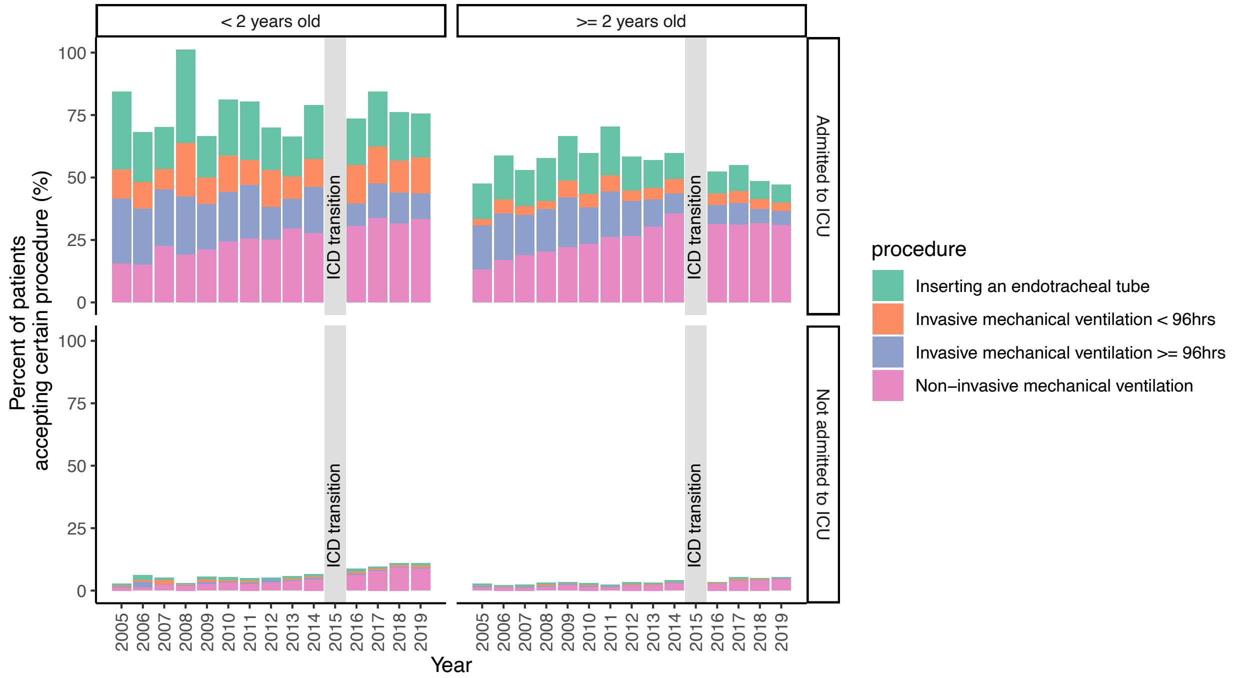


**Figure S8. Percent of RSV-associated hospitalized patients receiving certain medical support.**

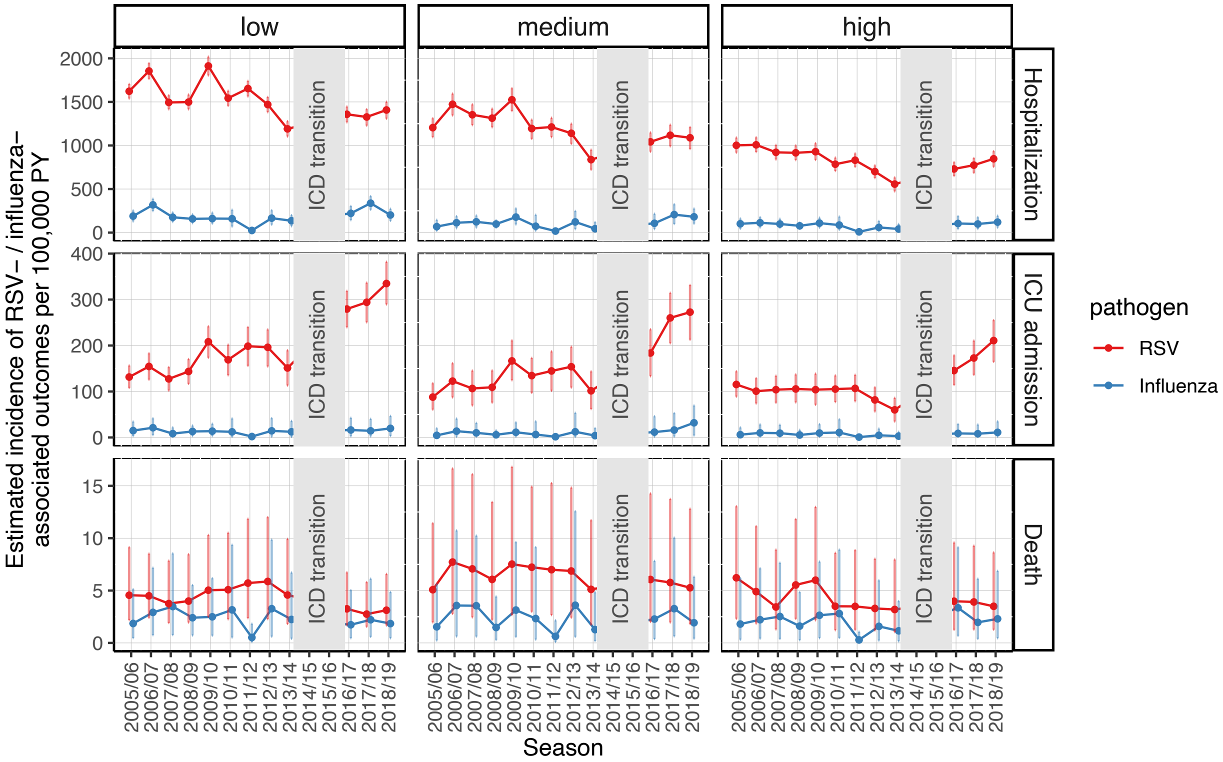


**Figure S9. Estimated incidence of RSV- and influenza-associated hospitalizations, ICU admissions, and deaths over time by SES group among infants under 2 years old, 2005-2019.** The dots represent the median estimates of incidence of hospitalizations, ICU admissions and deaths that are attributable to RSV or influenza per 100,000 person-years among age groups younger than 2 years old. The error bars indicate the 95% credible intervals of the estimated incidence.

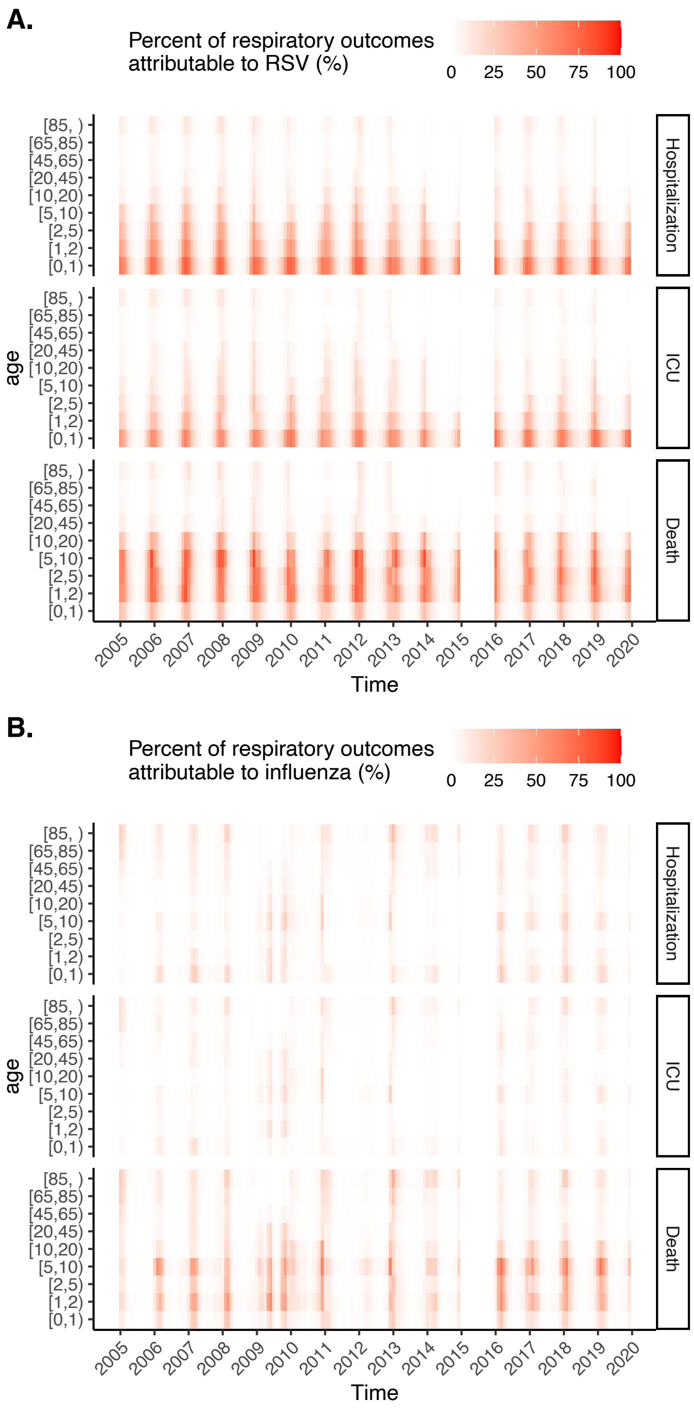


**Figure S10. Percent of respiratory outcomes attributable to (A) RSV and (B) influenza infection over time**. The shade of the rectangles indicates the median estimated percent of all-cause respiratory outcomes (hospitalizations, ICU admissions and deaths) that were attributable to RSV or influenza infection. The gap in 2015 indicates the transition from ICD-9-CM to ICD-10-CM code system.

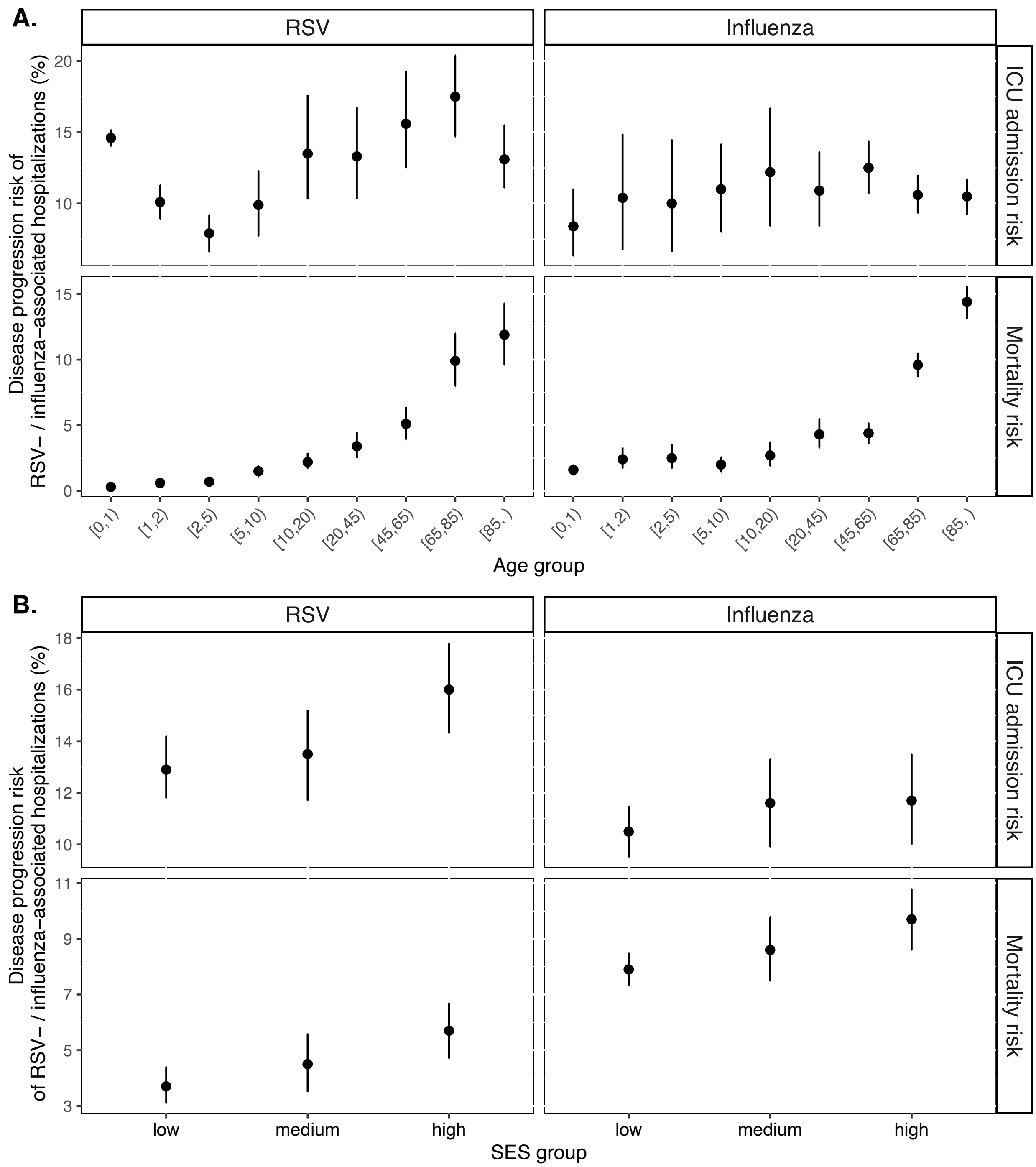


**Figure S11. Estimated ICU admission risk and mortality risk of RSV- and influenza-associated hospitalizations (A) by age and (B) by SES group, 2005-2019.** The dots and error bars indicate medians and 95% credible intervals of ICU admission risk and mortality risk. ICU admission risk is defined as the ratio between the incidence of virus-associated ICU admission and the incidence of virus-associated hospitalizations; the mortality risk is defined as the ratio between the incidence of virus-associated deaths and the incidence of virus-associated hospitalizations.

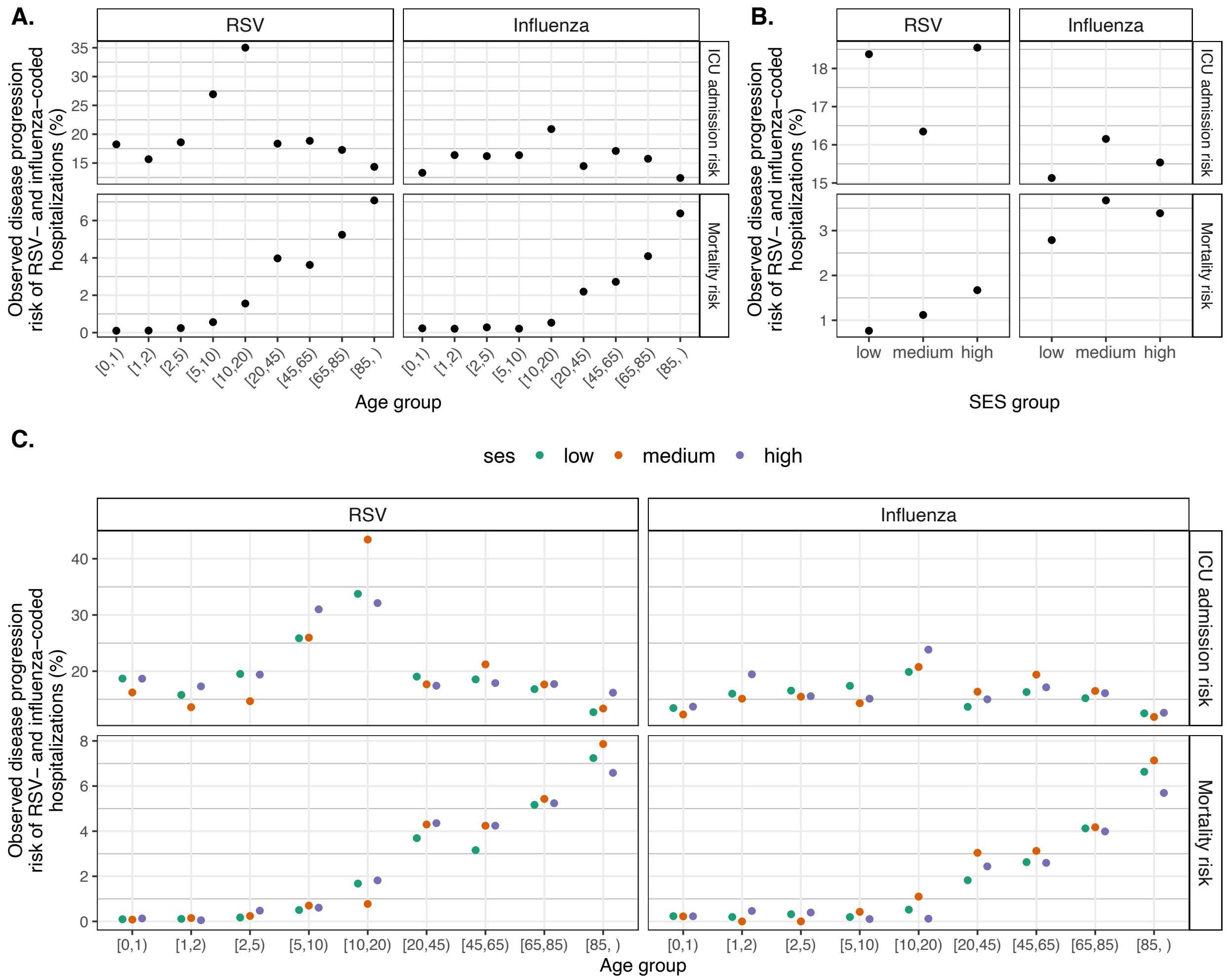


**Figure S12. Observed disease progression risk of RSV- and influenza-coded hospitalizations, 2005-2019.** (A) by age (B) by SES (C) by age and SES. The observed ICU admission risk was calculated as the ratio between the number of virus-coded ICU admissions and the number of virus-coded hospitalizations; the observed mortality risk was calculated as the number of virus-coded deaths and the number of virus-coded hospitalizations, aggregating all seasons from 2005/06 to 2018/19.

**Table S1 Estimated incidence of RSV-associated hospitalizations, ICU admissions, and deaths by season and age group. All numbers were presented as incidence per 100,000 person-years.**

| Outcome | Season | [0,1) | [1,2) | [2,5) | [5,10) | [10,20) | [20,45) | [45,65) | [65,85) | [85, ) |
|---|---|---|---|---|---|---|---|---|---|---|
| Hospitalization | **2005/2006** | 2083 (1989-2181) | 686 (618-744) | 239 (216-262) | 69 (57-81) | 22 (15-29) | 21 (15-28) | 36 (24-49) | 132 (97-171) | 769 (624-922) |
| | **2006/2007** | 2303 (2203-2404) | 808 (740-870) | 266 (240-291) | 77 (64-90) | 30 (22-38) | 23 (17-31) | 39 (28-51) | 160 (127-194) | 821 (693-952) |
| | **2007/2008** | 1923 (1837-2012) | 684 (620-745) | 249 (224-273) | 72 (59-84) | 30 (23-37) | 29 (22-36) | 60 (49-72) | 206 (171-240) | 900 (775-1027) |
| | **2008/2009** | 1984 (1898-2074) | 625 (556-691) | 232 (203-258) | 45 (34-58) | 24 (17-31) | 31 (24-39) | 57 (45-70) | 216 (181-252) | 798 (669-924) |
| | **2009/2010** | 2453 (2341-2567) | 713 (628-796) | 234 (195-271) | 38 (23-55) | 15 (8-23) | 20 (13-28) | 27 (18-40) | 119 (89-154) | 470 (347-609) |
| | **2010/2011** | 1894 (1800-1986) | 653 (587-715) | 251 (220-279) | 67 (54-80) | 29 (22-37) | 23 (17-30) | 35 (24-49) | 63 (37-96) | 402 (266-555) |
| | **2011/2012** | 2058 (1959-2153) | 635 (572-696) | 256 (224-286) | 68 (54-83) | 36 (27-46) | 28 (20-37) | 69 (53-84) | 200 (162-240) | 721 (579-875) |
| | **2012/2013** | 1912 (1809-2006) | 514 (458-570) | 193 (165-221) | 60 (45-76) | 25 (17-34) | 18 (12-25) | 41 (30-54) | 116 (94-141) | 393 (311-491) |
| | **2013/2014** | 1490 (1385-1589) | 430 (375-485) | 143 (117-169) | 40 (29-52) | 14 (8-21) | 10 (6-15) | 10 (5-19) | 15 (9-26) | 75 (41-128) |
| | **2016/2017** | 1761 (1664-1859) | 514 (447-575) | 163 (138-188) | 31 (20-44) | 27 (19-37) | 40 (30-50) | 35 (23-49) | 145 (111-178) | 623 (502-746) |
| | **2017/2018** | 1733 (1626-1835) | 561 (488-629) | 206 (178-233) | 44 (32-59) | 25 (17-35) | 38 (27-49) | 35 (23-49) | 140 (107-174) | 467 (352-593) |
| | **2018/2019** | 1793 (1691-1896) | 617 (537-692) | 216 (186-245) | 48 (34-63) | 20 (13-28) | 25 (18-33) | 24 (17-32) | 87 (71-107) | 204 (142-279) |
| ICU admission | **2005/2006** | 197 (170-226) | 41 (25-59) | 11 (6-18) | 3 (1-6) | 2 (1-4) | 3 (1-4) | 4 (1-8) | 23 (9-39) | 115 (68-165) |
| | **2006/2007** | 212 (180-245) | 55 (37-76) | 14 (7-22) | 4 (1-8) | 3 (1-6) | 3 (1-5) | 5 (3-9) | 30 (15-46) | 132 (83-182) |
| | **2007/2008** | 187 (158-217) | 43 (26-63) | 18 (11-26) | 5 (2-9) | 4 (2-6) | 4 (2-6) | 10 (6-14) | 38 (22-54) | 138 (87-188) |
| | **2008/2009** | 216 (186-246) | 35 (17-56) | 17 (9-26) | 4 (1-8) | 3 (1-6) | 4 (2-7) | 8 (4-13) | 36 (20-53) | 100 (56-149) |
| | **2009/2010** | 286 (249-323) | 60 (34-86) | 21 (10-32) | 4 (1-9) | 2 (0-4) | 3 (1-5) | 4 (1-8) | 20 (9-33) | 48 (17-92) |
| | **2010/2011** | 230 (196-266) | 59 (37-84) | 18 (9-28) | 7 (3-12) | 4 (2-7) | 4 (2-6) | 6 (2-12) | 7 (2-18) | 44 (14-89) |
| | **2011/2012** | 261 (219-303) | 64 (38-92) | 18 (10-29) | 8 (4-14) | 5 (2-8) | 4 (2-6) | 13 (7-19) | 31 (18-47) | 77 (36-131) |
| | **2012/2013** | 260 (217-300) | 58 (32-85) | 14 (6-24) | 7 (3-13) | 4 (2-7) | 2 (1-5) | 7 (4-12) | 18 (10-27) | 41 (17-77) |
| | **2013/2014** | 192 (150-230) | 45 (21-72) | 7 (2-16) | 4 (1-8) | 3 (1-5) | 1 (0-3) | 1 (0-3) | 1 (0-4) | 8 (2-23) |
| | **2016/2017** | 367 (326-410) | 86 (60-114) | 18 (9-28) | 4 (1-8) | 3 (1-6) | 5 (2-8) | 5 (2-10) | 22 (10-34) | 73 (35-116) |
| | **2017/2018** | 417 (370-462) | 97 (67-127) | 25 (13-36) | 7 (3-12) | 4 (2-7) | 5 (2-9) | 5 (2-11) | 28 (16-42) | 65 (27-111) |
| | **2018/2019** | 493 (442-543) | 90 (58-125) | 25 (13-39) | 8 (3-13) | 3 (2-6) | 3 (2-6) | 4 (1-7) | 23 (15-31) | 32 (12-59) |
| Death | **2005/2006** | 6.6 (3.4-12) | 3.8 (1.7-7) | 1.4 (0.7-2.6) | 0.7 (0.3-1.4) | 0.5 (0.2-1) | 0.5 (0.1-1) | 1.6 (0.5-3.6) | 10.1 (3.1-20.5) | 90 (38.7-146.1) |
| | **2006/2007** | 6.7 (3.6-12) | 3.9 (1.7-7.3) | 1.5 (0.7-2.8) | 0.9 (0.4-1.7) | 0.7 (0.2-1.2) | 0.4 (0.1-0.9) | 1.5 (0.5-3.3) | 18.3 (8.7-29.1) | 119.4 (74-168.4) |
| | **2007/2008** | 6 (3.1-11.3) | 2.8 (1.2-5.7) | 1.3 (0.5-2.6) | 1.2 (0.5-2) | 0.7 (0.2-1.2) | 0.7 (0.3-1.2) | 3.2 (1.6-5.1) | 22.1 (12-33.2) | 117.4 (76.8-163.9) |
| | **2008/2009** | 6 (3-11.6) | 3.7 (1.6-7.1) | 1.5 (0.6-3) | 0.8 (0.3-1.5) | 0.4 (0.1-0.9) | 0.7 (0.3-1.4) | 4.2 (2-6.8) | 23.6 (13-34.2) | 101 (57.4-148.5) |
| | **2009/2010** | 7.7 (3.9-14.2) | 4 (1.8-7.8) | 1.6 (0.6-3.1) | 0.5 (0.1-1.1) | 0.3 (0.1-0.8) | 0.4 (0.1-1.1) | 2 (0.7-4.2) | 13.6 (7.1-21.6) | 55.4 (28-97.7) |
| | **2010/2011** | 6.5 (3.1-12.7) | 3.8 (1.7-7.3) | 1.1 (0.5-2.2) | 0.7 (0.3-1.4) | 0.3 (0.1-0.8) | 0.5 (0.2-1.1) | 1.7 (0.5-3.6) | 2.6 (0.5-8.1) | 29 (7.1-68.4) |
| | **2011/2012** | 7.4 (3.5-14.5) | 3.5 (1.5-6.9) | 1.5 (0.6-3) | 0.8 (0.3-1.6) | 0.4 (0.2-0.9) | 0.6 (0.2-1.3) | 3.9 (1.8-6.5) | 18.1 (10.4-27.4) | 91.8 (48.5-144.3) |
| | **2012/2013** | 7.6 (3.3-14.7) | 3.5 (1.5-6.6) | 1.3 (0.5-2.7) | 1 (0.5-1.9) | 0.7 (0.3-1.2) | 0.4 (0.1-1) | 2.5 (1.1-4.4) | 11.2 (6.3-16.9) | 50.5 (26.7-78.7) |
| | **2013/2014** | 5.8 (2.5-11.8) | 3 (1.2-6.2) | 1 (0.4-2.3) | 0.8 (0.3-1.6) | 0.5 (0.2-1) | 0.3 (0.1-0.7) | 0.4 (0.1-1.2) | 0.5 (0.1-1.7) | 2.6 (0.6-9.4) |
| | **2016/2017** | 5.6 (2.9-10.7) | 2.5 (1-5.5) | 1.2 (0.5-2.3) | 0.6 (0.2-1.3) | 0.6 (0.2-1.2) | 2.3 (0.9-3.9) | 0.9 (0.2-2.2) | 13.2 (6.9-20.5) | 72.7 (38.9-107.3) |
| | **2017/2018** | 5.3 (2.7-9.6) | 2.2 (1-4.8) | 1.6 (0.7-3.1) | 0.5 (0.2-1.2) | 0.7 (0.2-1.4) | 1.9 (0.6-3.5) | 1 (0.3-2.6) | 12.6 (6.5-20.3) | 39.5 (15.7-70.1) |
| | **2018/2019** | 4.7 (2.5-8.9) | 2.7 (1.2-5.9) | 1.6 (0.7-3.1) | 0.5 (0.2-1.2) | 0.5 (0.2-1.1) | 0.9 (0.3-1.9) | 0.9 (0.2-2.1) | 11.1 (6.8-15.8) | 13.6 (3.3-32.5) |

**Table S2 Estimated incidence of Influenza-associated hospitalizations, ICU admissions, and deaths by season and age group. All numbers were presented as incidence per 100,000 person-years.**

| Outcome | Season | [0,1) | [1,2) | [2,5) | [5,10) | [10,20) | [20,45) | [45,65) | [65,85) | [85, ) |
|---|---|---|---|---|---|---|---|---|---|---|
| Hospitalization | 2005/2006 | 248 (177-319) | 40 (13-83) | 16 (4-35) | 16 (8-25) | 3 (1-6) | 6 (3-11) | 44 (33-56) | 273 (241-307) | 750 (631-871) |
| | 2006/2007 | 348 (276-420) | 102 (50-155) | 14 (4-30) | 16 (7-28) | 4 (1-8) | 20 (14-27) | 79 (65-94) | 304 (269-341) | 931 (806-1071) |
| | 2007/2008 | 240 (178-305) | 47 (21-81) | 11 (4-24) | 10 (5-19) | 5 (2-9) | 21 (16-27) | 80 (69-92) | 321 (287-353) | 1339 (1222-1464) |
| | 2008/2009 | 155 (107-202) | 96 (58-136) | 32 (16-51) | 39 (29-49) | 19 (12-26) | 19 (14-24) | 37 (28-46) | 63 (45-85) | 166 (127-217) |
| | 2009/2010 | 184 (122-254) | 117 (68-178) | 49 (23-80) | 54 (38-69) | 28 (19-38) | 27 (20-34) | 51 (39-64) | 117 (87-149) | 354 (274-445) |
| | 2010/2011 | 198 (98-315) | 51 (18-116) | 26 (8-51) | 23 (13-37) | 17 (11-24) | 23 (14-33) | 76 (59-94) | 343 (290-393) | 1143 (943-1341) |
| | 2011/2012 | 30 (14-51) | 8 (3-19) | 4 (1-10) | 8 (5-12) | 4 (2-6) | 4 (3-6) | 8 (6-12) | 61 (51-71) | 244 (207-280) |
| | 2012/2013 | 227 (135-335) | 36 (14-77) | 17 (5-41) | 25 (16-37) | 12 (6-20) | 24 (16-32) | 77 (64-92) | 467 (423-509) | 1863 (1708-2017) |
| | 2013/2014 | 172 (99-248) | 19 (7-44) | 5 (2-11) | 5 (2-10) | 3 (1-7) | 7 (3-12) | 85 (72-99) | 232 (204-261) | 1217 (1094-1328) |
| | 2016/2017 | 273 (182-364) | 68 (29-129) | 14 (6-25) | 40 (25-55) | 14 (5-23) | 28 (18-38) | 81 (66-96) | 222 (186-263) | 905 (762-1042) |
| | 2017/2018 | 391 (303-478) | 115 (63-179) | 49 (30-70) | 50 (36-64) | 22 (14-30) | 40 (31-50) | 80 (66-93) | 300 (268-335) | 1446 (1326-1562) |
| | 2018/2019 | 272 (196-347) | 85 (28-146) | 16 (8-31) | 32 (20-45) | 10 (5-17) | 22 (14-30) | 77 (65-89) | 209 (182-238) | 757 (662-855) |
| ICU admission | 2005/2006 | 18 (5-40) | 3 (1-13) | 2 (0-7) | 1 (0-4) | 0 (0-1) | 0 (0-1) | 5 (1-8) | 43 (28-58) | 53 (22-92) |
| | 2006/2007 | 30 (11-54) | 4 (1-12) | 1 (0-5) | 1 (0-3) | 0 (0-1) | 3 (1-5) | 11 (7-16) | 24 (11-39) | 88 (47-131) |
| | 2007/2008 | 15 (5-32) | 3 (1-10) | 1 (0-5) | 1 (0-3) | 0 (0-2) | 2 (1-4) | 10 (6-15) | 29 (16-44) | 137 (90-185) |
| | 2008/2009 | 7 (2-17) | 13 (3-25) | 3 (1-8) | 4 (2-7) | 2 (1-4) | 4 (2-5) | 4 (2-8) | 6 (2-13) | 15 (8-28) |
| | 2009/2010 | 10 (2-25) | 15 (4-31) | 5 (1-11) | 6 (2-10) | 4 (1-7) | 5 (3-7) | 8 (4-13) | 13 (5-24) | 56 (33-85) |
| | 2010/2011 | 17 (3-50) | 5 (1-20) | 2 (0-6) | 2 (1-6) | 3 (1-5) | 2 (0-4) | 11 (5-18) | 46 (28-66) | 152 (88-223) |
| | 2011/2012 | 3 (0-9) | 1 (0-3) | 0 (0-1) | 1 (0-3) | 0 (0-1) | 1 (0-1) | 1 (0-2) | 5 (2-9) | 23 (12-36) |
| | 2012/2013 | 21 (5-54) | 3 (1-12) | 1 (0-5) | 4 (1-8) | 1 (0-3) | 2 (1-5) | 8 (3-13) | 42 (27-58) | 203 (148-262) |
| | 2013/2014 | 16 (3-42) | 2 (0-7) | 0 (0-1) | 0 (0-1) | 0 (0-1) | 1 (0-2) | 4 (1-8) | 17 (8-26) | 96 (51-142) |
| | 2016/2017 | 21 (5-51) | 7 (1-21) | 1 (0-4) | 3 (1-6) | 2 (0-4) | 3 (1-6) | 15 (9-20) | 29 (16-43) | 86 (40-137) |
| | 2017/2018 | 20 (5-49) | 7 (2-23) | 3 (1-9) | 3 (1-8) | 2 (0-5) | 2 (1-5) | 6 (2-11) | 24 (12-37) | 160 (114-209) |
| | 2018/2019 | 31 (10-60) | 9 (1-29) | 1 (0-5) | 6 (1-11) | 1 (0-2) | 2 (0-4) | 10 (5-15) | 20 (10-31) | 123 (83-163) |
| Death | 2005/2006 | 2.4 (0.8-6.1) | 1.3 (0.3-3.9) | 0.3 (0.1-1) | 0.5 (0.1-1.2) | 0.1 (0-0.5) | 0.2 (0.1-0.6) | 1.9 (0.6-3.7) | 25.3 (16.7-34.2) | 98.8 (55-144.2) |
| | 2006/2007 | 4.4 (1.4-9.4) | 1.5 (0.4-3.9) | 0.4 (0.1-1.2) | 0.6 (0.2-1.4) | 0.1 (0-0.4) | 0.6 (0.2-1.2) | 4.8 (2.8-7) | 32.2 (22.3-43) | 137.2 (90-184.4) |
| | 2007/2008 | 4.8 (1.5-10.3) | 1.8 (0.4-4.6) | 0.4 (0.1-1.3) | 0.3 (0.1-0.9) | 0.2 (0-0.6) | 0.7 (0.2-1.3) | 4.4 (2.4-6.4) | 36.5 (27.2-46.6) | 208.5 (164.5-254.2) |
| | 2008/2009 | 2.5 (0.8-6) | 1.7 (0.5-3.7) | 0.5 (0.2-1.3) | 0.3 (0.1-0.8) | 0.2 (0.1-0.5) | 0.9 (0.5-1.4) | 1.6 (0.6-2.9) | 4 (2.2-7.5) | 18.4 (12.8-28.1) |
| | 2009/2010 | 3.4 (1-7.9) | 2.2 (0.6-5.1) | 0.8 (0.2-1.9) | 0.5 (0.2-1.1) | 0.5 (0.1-1) | 1.1 (0.5-1.7) | 2.6 (0.9-4.6) | 11.7 (7.2-17.8) | 57.9 (37.8-82) |
| | 2010/2011 | 4.5 (1-11.5) | 1.6 (0.4-4.6) | 0.5 (0.1-1.6) | 0.5 (0.1-1.2) | 0.4 (0.1-1) | 0.6 (0.1-1.3) | 5.7 (2.9-8.7) | 46 (32.5-60.2) | 220 (156.2-284) |
| | 2011/2012 | 0.8 (0.2-2.2) | 0.2 (0-0.7) | 0.1 (0-0.3) | 0.2 (0-0.4) | 0.1 (0-0.2) | 0.1 (0-0.3) | 0.6 (0.3-1.2) | 7.9 (5.2-10.6) | 34.7 (22.1-47.2) |
| | 2012/2013 | 4.7 (1.1-12.3) | 1.4 (0.3-4.2) | 0.5 (0.1-1.7) | 0.5 (0.1-1.4) | 0.3 (0.1-0.9) | 0.7 (0.2-1.4) | 4.6 (2.5-7.2) | 50 (39-61) | 283.5 (229.3-338.2) |
| | 2013/2014 | 2.9 (0.7-7.9) | 0.8 (0.2-2.5) | 0.2 (0-0.7) | 0.3 (0-0.9) | 0.2 (0-0.6) | 0.3 (0.1-0.8) | 2.3 (1-4.1) | 20.3 (13.4-27.6) | 187.2 (146.8-225.7) |
| | 2016/2017 | 3.1 (1-7.1) | 1.7 (0.4-4.1) | 0.4 (0.1-1.3) | 0.6 (0.2-1.3) | 0.4 (0.1-1) | 1.6 (0.5-3.3) | 2.5 (0.9-4.4) | 19.9 (11.8-28.5) | 94.7 (60-134.6) |
| | 2017/2018 | 3.4 (1.1-8) | 1.5 (0.4-4) | 0.9 (0.2-2.1) | 0.6 (0.2-1.3) | 0.4 (0.1-0.9) | 2.3 (1-3.8) | 2.1 (0.7-3.8) | 16.5 (9-24.2) | 158.9 (124.6-193.8) |
| | 2018/2019 | 2.7 (0.9-6.3) | 1.4 (0.4-3.7) | 0.6 (0.1-1.6) | 0.7 (0.2-1.3) | 0.3 (0.1-0.8) | 1.1 (0.4-2.3) | 2.2 (0.8-3.8) | 14 (8.1-20.4) | 94.7 (67.7-123.9) |
